# Gut microbiota dysbiosis in adrenoleukodystrophy: a new piece in the puzzle?

**DOI:** 10.64898/2026.10.19.26365873

**Authors:** Lorenzo Torreni, Agatha Schlüter, Sara Marti, Montserrat Ruiz, Valentina Vélez-Santamaria, Agustí Rodríguez-Palmero, Ayelen Artuch, Cristina Guilera, Angela Arias, Fanny Mochel, Sergio Aguilera-Albesa, Veronica Cantarin, Carlos Casasnovas, Rafael Artuch, Wolfgang Köhler, Aurora Pujol, Stéphane Fourcade

**Author notes:** Correspondence to: Stéphane Fourcade and Aurora Pujol, Neurometabolic Diseases Laboratory, IDIBELL, Gran Via, 199, 08908 L’Hospitalet de Llobregat, Barcelona, Spain.

## Abstract

X-linked adrenoleukodystrophy (X-ALD) is the most common monogenic disorder affecting brain white matter, presenting a broad clinical spectrum from slowly progressive adrenomyeloneuropathy (AMN) to rapidly advancing cerebral demyelination. Factors influencing the variable disease expressivity remain poorly understood. We conducted shotgun metagenomic profiling of fecal samples from X-ALD patients and integrated these data with targeted plasma metabolomic analyses of amino acids and key metabolic intermediates. We identified distinct microbiome-associated metabolic signatures across X-ALD phenotypes. Notably, *Bacteroides cellulosilyticus* and *Helicobacter apodemus* abundance were significantly altered in AMN compared with childhood CALD patients. In parallel, X-ALD patients exhibited evidence of microbial metabolic remodeling characterized by altered amino acid metabolism, reduced folate biosynthetic potential, and enrichment of stress-adaptive pathways, including redox-associated functions, glyoxylate shunt activity, lipid A modification, lipopolysaccharide assembly, biofilm adhesin biosynthesis, and curli fiber production. Consistent with these microbial changes, plasma levels of branched-chain amino acids and methionine, as well as serum folate concentrations, were significantly reduced. Importantly, methionine levels showed an inverse correlation with neurofilament light chain (NfL) concentrations in patients with cerebral disease. Our findings reveal a previously unrecognized link between gut microbial metabolic capacity and systemic metabolic alterations in X-ALD. Our results reinforce previous evidence implicating methionine metabolism in disease severity. Collectively, these data support a modulatory role of the microbiota–gut–brain axis in neuroinflammatory and mitochondrial-related processes underlying X-ALD progression and highlight new opportunities for biomarker development and microbiome-informed therapeutic interventions.

## INTRODUCTION

X-linked adrenoleukodystrophy (X-ALD, OMIM 300100) is one of the most prevalent leukodystrophy, affecting roughly 1 in 14,700 newborns ^1,2^. Mutations in the *ABCD1* gene, located on Xq28, cause the disease. This gene encodes the ATP-binding cassette half-transporter ALDP (adrenoleukodystrophy protein) localized on peroxisomal membranes ^1–3^. Dysfunction of this transporter leads to the buildup of its preferred substrates, very long-chain fatty acids (VLCFAs) with straight chains, particularly C26:0, in plasma and tissues. The brain’s white matter, spinal cord, adrenal cortex, and testicular Leydig cells are most severely affected ^1–3^. The precise mechanisms by which these fatty acids exert neurotoxicity remain unclear. Our research, along with others’, suggests that excess VLCFAs disrupt the mitochondrial transport chain, leading to free radical production ^4^ and redox imbalance ^5–8^.

X-ALD patients exhibit three major neurological presentations ^1–3,9,10^: 1) cerebral adrenoleukodystrophy (CALD) in childhood, that affects approximately 35% of male X-ALD patients. It manifests early (3-10 years old, peaking at 7) and progresses rapidly, leading to severe disability within 6 months to 2 years. It is characterized by pronounced inflammatory demyelination in the cerebral white matter; 2) adrenomyeloneuropathy (AMN): typically appearing after 30 years with a gradual progression over decades, AMN presents with sensory ataxia, progressive spastic paraparesis, leg pain, sphincter dysfunction, and impotence due to progressive damage to the spinal cord and peripheral nerves; 3) adult CALD: occurring in likely more than 50% of men, adult CALD combines an AMN-like clinical course with cerebral inflammatory demyelination. Additionally, nearly 80% of male patients experience signs and symptoms of adrenocortical insufficiency, with or without neurological involvement. Allogeneic hematopoietic stem cell transplantation (HSCT) can halt disease progression of CALD if performed early, before extensive demyelination occurs ^11–14^. Redox imbalance appears linked to inflammation since a phase II/III clinical trial with high-dose antioxidants normalized oxidative damage and inflammatory markers, and improved locomotor tests in AMN patients ^15^. Leriglitazone, a natural derivative of the PPARy agonist pioglitazone which demonstrated efficacy in X-ALD mouse models ^16^, may offer an alternative therapeutic option at the early stages of CALD for patients who are not candidates for HSCT ^17–19^.

The vast spectrum of clinical variations observed within families suggests the influence of unknown genetic, epigenetic, or environmental modifiers ^3^. Currently, VLCFA blood level measurement holds diagnostic value only and there is still not a clinically approved prognostic biomarker for X-ALD’s natural history ^10^, although levels of lyso-phosphatidylcholine C26:0 has recently been shown to correlate with disease severity ^20^. The human gut harbors a vast array of commensal microorganisms, collectively known as the gut microbiota. We have co-evolved with this microbial ecosystem, and its immense genetic potential is thought to influence virtually all human physiological processes, including metabolism and immune homeostasis: due to its profound impact, the gut microbiota has been dubbed the “forgotten organ” ^21^. Notably, the gut microbiota plays a crucial role in bidirectional communication between the gut and the central nervous system (CNS) via immunological, humoral, and neuronal signals, forming the microbiota-gut-brain axis (MGBA) ^21–23^. Disruptions in this communication are implicated in various neurological diseases, including Parkinson’s disease (PD), Alzheimer’s disease (AD), and multiple sclerosis (MS) ^23–25^ where the MGBA may influence disease risk, activity, and progression ^25–28^. Moreover, gut microbiota alterations are linked not only to neurological disorders but also to metabolic and autoimmune diseases ^29,30^.

The intricate relationship between the host and its microbiome in regulating neurological, immune, and metabolic functions in health and disease suggests that gut microbiota could be an important environmental factor in X-ALD pathogenesis. In this context, gut microbiota dysbiosis emerges as a potential driver of the inflammatory phenotype proposed in the ’three-hit’ hypothesis of X-ALD ^31^, consistent with observations of microbiome alterations reported in X-ALD mouse models^32^. Furthermore, investigating the microbiome and its associated metabolites could offer a rationale for microbe-based therapies and the implementation of microbial derived biomarkers for diagnostic and/or prognostic purposes. We used shotgun metagenomic analysis of fecal samples and targeted metabolomic analysis in plasma from X-ALD patients to characterize the taxonomic composition of the gut microbiota, its functional potential and a metabolite signature in these patients.

## MATERIAL AND METHODS

### Participants and plasma collection

Plasma was collected from 68 male X-ALD patients (17 with childhood CALD, mean age 9 years; 43 with AMN, mean age 45.6 years; 8 with adult CALD, mean age 38.8 years) and 64 age-matched controls (35 children, mean age 10.6 years; 29 adults, mean age 37.9 years). Peripheral venous blood was collected in EDTA-containing tubes, centrifuged, and the plasma obtained was immediately frozen at -80°C until analysis.

### Participants and stool collection

Stool samples were collected from 32 male adult X-ALD patients (30 with AMN and 2 adult CALD, mean age 44.6 years), 6 male childhood CALD patients (mean age 9 years), and 48 male controls (19 children, mean age 8.7 years; 29 adults, mean age 43 years) using the OMNIgene-GUT™ kit (DNA Genotek).

### DNA extraction

DNA was obtained from stool samples using the Qiagen Power Fecal Pro DNA kit according to the manufacturer’s instructions and protocols. DNA quality was evaluated visually via gel electrophoresis and quantified using a Qubit 3.0 fluorometer (Thermo-Fischer, Waltham, MA, USA).

The use of human samples was approved by the Clinical Research Ethics Committee of the Bellvitge University Hospital (AC130/10, PR170/18, PR328/20 and PR261/22).

### Shotgun metagenomic

Libraries were prepared using an Illumina Nextera library preparation kit with an in-house protocol (Illumina, San Diego, CA, USA). Paired-end sequencing (150 bp x 2) was done on a NextSeq 500 in medium-output mode. Shotgun metagenomic sequence reads were processed with the Sunbeam pipeline. Initial quality evaluation was done using FastQC v0.11.5 (http://www.bioinformatics.babraham.ac.uk/projects/fastqc). Processing took part in four steps: adapter removal, read trimming, low-complexity-reads removal, and host-sequence removals. Adapter removal was done using cutadapt v2.6 ^33^. Trimming was done with Trimmomatic v0.36 ^34^ using custom parameters (LEADING:3 TRAILING:3 SLIDINGWINDOW:4:15 MINLEN:36). Low-complexity sequences were detected with Komplexity v0.3.6 ^35^. High-quality reads were mapped to the human genome (Genome Reference Consortium Human Reference 37) and those that mapped to it were with at least 50% similarity across 60% of the read length were removed.

### Targeted amino acid analysis

The amino acid standard solutions and plasma samples from X-ALD patients and controls were analyzed to determine amino acid levels using a Waters ACQUITY UPLC H-class instrument equipped with a reversed-phase C-18 column. Detection was performed with a Waters Xevo TQD triple-quadrupole mass spectrometer using positive electrospray ionization in the multiple reaction monitoring (MRM). Further details on sample treatment, chromatographic and mass spectrometry conditions can be found in ^36^. This procedure is accredited following the ISO15189 norm and is under external quality control assessment (www.erndim.org).

### Neurofilament Light Chain and Glial Fibrillary Acidic Protein Determination

Analysis of plasma samples from X-ALD patients was conducted at the Vall d’Hebron Institut de recerca (VHIR) using the Single Molecule Array (SiMoA™) HD-1 Analyzer (Quanterix Corporation), which runs ultrasensitive paramagnetic bead-based enzyme-linked immunosorbent assays ^37^. Plasma concentration determinations of neurofilament-light chain (NfL) and glial fibrillary acidic protein (GFAP) were performed according to the manufacturer’s instructions and protocols with appropriate calibrators and internal controls (www.quanterix.com/products-technology/assays).

### Statistical analysis

No formal sample size calculation was performed, and missing data were not imputed. We minimized potential sources of bias by including untreated participants, ensuring no significant age differences between groups, and applying standardized protocols to reduce technical batch effects; however, potential confounding due to geographic heterogeneity, given that X-ALD samples were obtained from Spain, Germany and France, and controls from Spain, France and the Netherlands, as well as unmeasured factors such as diet, cannot be fully excluded. For the shotgun metagenomic analysis, after quality control and removal of human-derived reads, taxonomic classification of the remaining sequences was performed using Kraken2 with the PlusPF database ^38^. For functional profiling, high-quality filtered reads were aligned against the SEED database using a translated homology search. Subsequently, these reads were annotated to functional levels 1-3 (Subsystems) using Super-Focus ^39^. The vegan package (https://CRAN.R-project.org/package=vegan) was employed for ordination analysis (beta diversity) of taxonomic profiles, comparing species-level taxonomic profiles using Bray-Curtis dissimilarities. Non-metric multidimensional scaling (NMDS) was used to visualize sample distribution based on these dissimilarities. The Shannon diversity index was employed to assess alpha diversity, integrating species presence and proportional abundance in each microbial community. Differential abundance analysis was performed using the DESeq2 R package ^40^: we employed a negative binomial model to test for significant differences in abundance of taxonomic features and functional categories. The analysis aimed to identify differences between groups and disease status while controlling for age differences (sequential model). p-values were calculated using the likelihood ratio test (LRT) with an adjusted threshold of 0.05. All statistical analyses were conducted using R software (https://www.R-project.org). To assess the effect of Genotype (control vs. X-ALD) and Age (child vs. adult) on bacterial abundance, a two-way analysis of variance (2-way ANOVA) was applied when data satisfied the assumptions of normality and homoscedasticity. For datasets that did not meet these parametric assumptions, the non-parametric Scheirer–Ray–Hare test, an extension of the Kruskal–Wallis test for two factors, was used as a robust alternative. Post-hoc pairwise comparisons among the four experimental groups (child control, childhood CALD, adult control, and AMN) were conducted using Wilcoxon rank-sum tests. The resulting p-values from both the ANOVA and Wilcoxon analyses were corrected for multiple testing using the Benjamini–Hochberg false discovery rate (FDR) procedure.

Serum folate (folic acid/folate) concentrations were obtained from routine clinical blood analyses performed at participating hospitals in Spain and Serbia. Measurements were reported in either ng/mL or nmol/L depending on the laboratory and were interpreted according to each center’s validated reference ranges. Folate deficiency was defined using the lower limit of the corresponding hospital-specific reference interval. To account for inter-laboratory variability, folate status was analyzed as a binary variable (deficient vs. non-deficient). The observed prevalence of folate deficiency was compared with an expected prevalence of 10% in the general population using an exact one-sample binomial test. The distribution of amino acids, NfL and GFAP levels in plasma from controls, AMN, childhood and adult CALD groups was assessed using the Shapiro-Wilk test. Outliers were identified using the Rout test and excluded from subsequent analyses. Unpaired t-tests were used to compare plasma amino acid levels between the child control group and the childhood CALD group, provided the data distribution was normal. Otherwise, the non-parametric Mann-Whitney test was employed. For the adult groups (controls, AMN, and adult CALD), one-way ANOVA was used for normally distributed data, while the Kruskal-Wallis test was applied for non-normal data. The threshold for statistical significance (p-value) was set at 0.05. Spearman’s rank correlation coefficient was calculated to assess the correlation between amino acids levels and NfL or GFAP levels. The p-value threshold for correlation analysis was set at 0.05.

We calculated the area under the curve (AUC) of the receiver operating characteristic (ROC) curve of the amino acids that were significantly different between X-ALD patients and controls to evaluate their diagnostic accuracy. The confidence interval was set at 95% and the p-value threshold was set at 0.05.

Statistical analyses and graphs for amino acids levels comparison between groups, their ROC curves and their correlation with NfL and GFAP levels were performed using GraphPad Prism version 9.4.1. Box plots illustrating significant differences in bacterial taxa or functions identified by shotgun metagenomic analysis were also created using GraphPad Prism version 9.4.1. All other figures were generated using R software ^41^.

### Ethics approval and consent to participate

The use of human samples in this study was approved by the Clinical Research Ethics Committee of the Bellvitge University Hospital under the protocols AC130/10, PR170/18, PR328/20, and PR261/22. All participants or their legal guardians provided informed consent in accordance with the Declaration of Helsinki.

## RESULTS

### The gut microbiota is altered in X-ALD patients

We conducted an analysis of gut microbiota composition using shotgun metagenomics. At the phylum level, the fecal bacterial profiles of both X-ALD patients and controls were dominated by Bacteroidetes and Firmicutes (Fig. 1A). At the genus level, *Bacteroides*, *Faecalibacterium*, and *Phocaeicola* were the most abundant taxa (Fig. 1B). Analysis of beta diversity (species diversity between samples) using Bray-Curtis dissimilarities ^42^ did not reveal a clear separation between the childhood and adult CALD, AMN, and their respective control groups (Fig. 1C). Similarly, alpha diversity, measured using Shannon’s diversity index ^43,44^, did not differ significantly across groups (Fig. 1D). A sequential model, however, suggested a potential age-related effect. Notably, no significant difference in Shannon diversity was observed between patients with the childhood CALD and those with AMN. Differential abundance analysis between childhood CALD and AMN patients identified 60 bacterial species, with 34 showing increased abundance in childhood CALD patients (Fig. 2A and Fig. S1) using 2-way ANOVA or the non-parametric Scheirer–Ray–Hare test (for non-normal data), followed by Wilcoxon rank-sum post-hoc comparisons with Benjamini–Hochberg FDR correction. Most of the identified species are not currently known to be either pathogenic or protective in any disease context. Two notable exceptions are *Helicobacter apodemus* (*H. apodemus*) and *Bacteroides cellulosilyticus* (*B. cellulosilyticus*). *H. apodemus*, which is reduced in AMN patients, has been implicated in intestinal inflammation by disrupting innate lymphoid cell (ILC) homeostasis, particularly via suppression of ILC3, and by promoting the production of pro-inflammatory cytokines ^45–47^. Accordingly, reduced *H. apodemus* abundance in AMN patients may be protective. In contrast, *B. cellulosilyticus*, which is increased in AMN patients, has been reported to exert anti-inflammatory effects through the induction of IL-10 ^48^, consistent with the elevated IL-10 expression observed in these patients ^15,49^.

**Figure 1.**
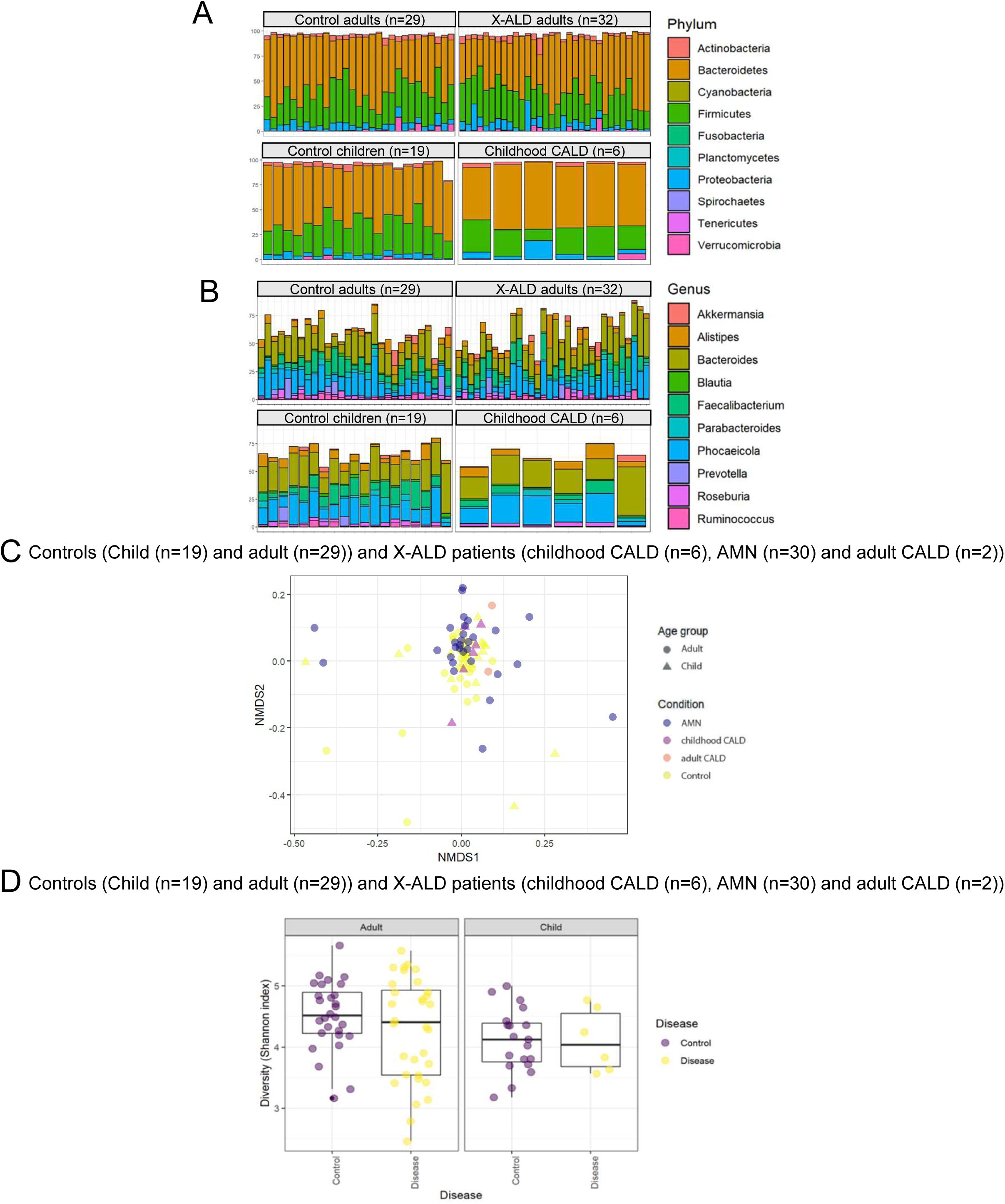
X-ALD gut microbiota taxonomic profile. (A, B) Bar charts showing the relative abundances (%) of the ten most abundant bacteria phila (A) and genera (B) in X-ALD (adult CALD (n=2), childhood CALD (n=6), AMN (n=30) and controls (n= 19 child and n=29 adult) stool samples. (C) Bidimensional PCA corresponding to the Bray-Curtis dissimilarity index (beta diversity) in X-ALD and controls samples (AMN (n=30), childhood CALD (n=6), adult CALD (n=2), child control (n=19), and adult control (n=29). Samples were visualized using non-metric multidimensional scaling (NMDS). (D) Box plots showing the Shannon index distribution (alpha diversity) in adults and child X-ALD patients and Controls.

**Figure 2.**
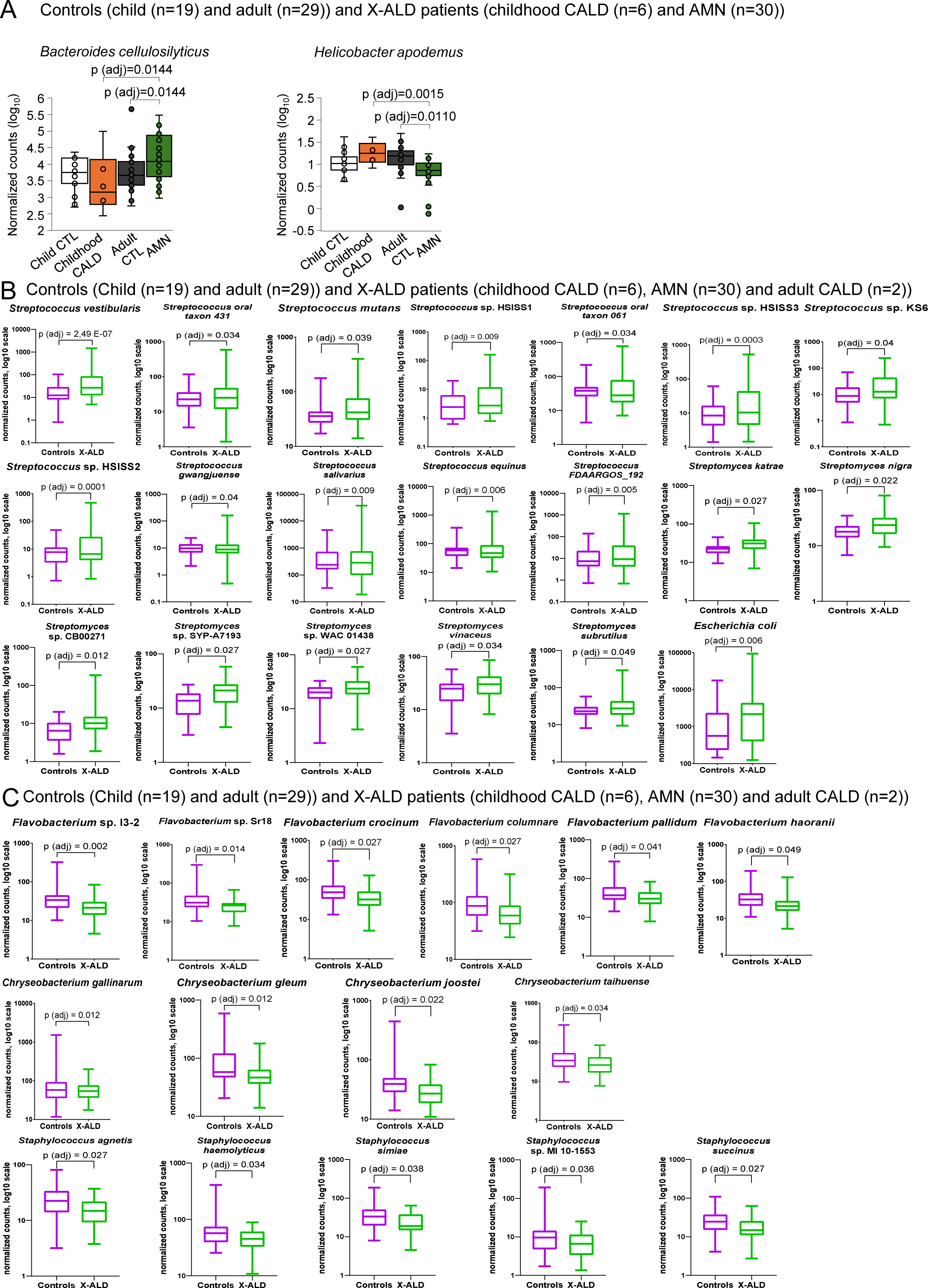
The most consistent alterations at species level in X-ALD microbiota. (A) Box plots showing the differential abundances (normalised read counts on log10 scale) of several species in childhood CALD (n=6), AMN (n=30) respect to healthy patients (n= 19 child and n=29 adult). Whiskers from minimum to maximum, median abundances and interquartile ranges have been indicated in the plots. We looked for differences between among groups, and also for difference in disease status after controlling for age differences. Statistical significance was assessed using 2-way ANOVA or the non-parametric Scheirer–Ray–Hare test (for non-normal data), followed by Wilcoxon rank-sum post-hoc comparisons with Benjamini–Hochberg FDR correction. Box plots showing the differential abundances (normalised read counts on log10 scale) of several species of *Streptococcus* spp and *Streptomyces* spp, and *Escherichia coli* (B) *Flavobacterium* spp, *Chryseobacterium* spp and *Staphylococcus* spp (C) in X-ALD patients (n=38) respect to controls (n=48). Negative binomial models were used for differential abundance testing of taxonomic features. We looked for differences between among groups, and also for difference in disease status after controlling for age differences. p-values were calculated with LRT test with an adjusted p-value cut off = 0.05. Whiskers from minimum to maximum, median abundances and interquartile ranges have been indicated in the plots.

We next proceeded to compare the combined X-ALD group (childhood and adult CALD + AMN) with the combined control group (adult + child), controlling for age to identify microbial signatures associated with disease status, using Likelihood Ratio Test (LRT). This analysis revealed 226 significantly different taxa and 144 differentially abundant functions in X-ALD patients compared to controls (see Supplementary Material ES1 and ES2). Among the most consistent taxonomic alterations, we observed increased abundance of *Streptococcus* spp, and *Streptomyces* spp. and *Escherichia coli* (Fig. 2B), and decreased abundance of *Flavobacterium* spp., *Chryseobacterium* spp., and *Staphylococcus* spp. (Fig. 2C).

From a functional perspective, two main patterns emerged: a) Amino acid metabolism: X-ALD patients exhibited a reduction in bacterial functions involved in the biosynthesis of several amino acids, including branched-chain amino acids (BCAAs: isoleucine, leucine, and valine), as well as glutamine, glutamate, aspartate, asparagine, serine, alanine, cysteine, proline, and methionine. This was accompanied by an increased abundance of lysine degradation pathways (Fig. 3); b) Bacterial adaptation to environmental stress. Metagenomic analysis revealed a reduction in functional categories related to tRNA aminoacylation capacity, specifically for tryptophan, together with the decreased abundance of ct⁶A nucleoside biosynthesis pathways, suggesting a disruption in the translational machinery of the gut microbiota in childhood CALD compared to AMN patients. These impairments may compromise the fidelity and efficiency of protein synthesis, particularly under stress conditions where accurate decoding is critical. We observed a decrease in At1g01770-like genes, which encode proteins containing a conserved but uncharacterized stress-related protein DUF1446 domain, potentially involved in redox or environmental adaptation. (Fig. 4A). We also observed a reduced functional potential for microbial vitamin B6 and folate biosynthesis in X-ALD patients compared with controls (Fig. 4B). We then measured serum folate (folic acid/folate) as part of routine clinical blood analyses in a subset of patients from several Spanish and Serbian hospitals, including AMN (n = 14), childhood CALD (n = 8), and adult CALD (n = 6). Overall, 11 of 28 patients met criteria for folate deficiency (5 AMN, 2 childhood CALD, and 4 adult CALD) based on hospital-specific laboratory reference ranges (Table 1). A one-sample binomial test demonstrated that the observed prevalence of folate deficiency was significantly higher than expected in the general population (*p* < 0.0001). These findings are consistent with metagenomic data indicating reduced microbial folate biosynthetic potential in X-ALD patients (Fig. 4B). Finally, we also detected a consistent increase in pathways associated with oxidative stress regulation, encompassing genes encoding key redox regulators (e.g., OxyR, SoxR, PerR) and antioxidant enzymes (such as catalases, peroxidases, and superoxide dismutases), as well as genes belonging to the universal stress protein (USP) family, which are typically involved in bacterial survival under prolonged environmental stress. We detected additional enrichment for pathways related to the glyoxylate bypass, lipid A modification, lipopolysaccharide assembly, biofilm adhesin biosynthesis, and Curli fiber production, amyloid fibers that form part of bacterial biofilms and are structurally similar to human amyloids (Fig. 4C).

**Figure 3.**
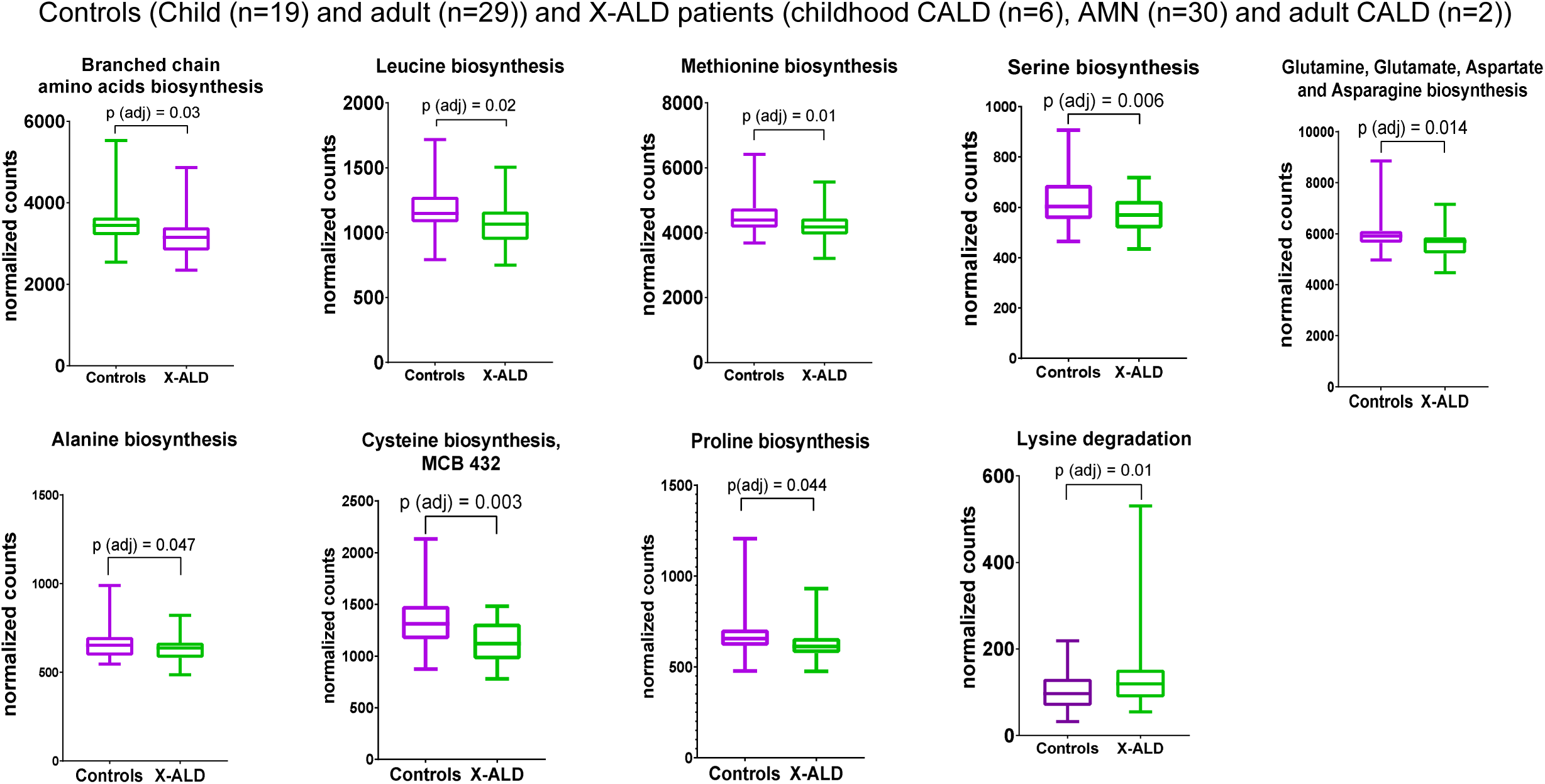
Microbiota functional features of altered amino acids metabolism in X-ALD patients. Box plots showing the differential abundances (normalised read counts) of functions implicated in amino acid metabolism in the gut microbiota of X-ALD (n=38) patients compared to controls (n=48). Negative binomial models were used for differential abundance testing of subsystem level 3 (function) features. p-values were calculated with LRT test with an adjusted p-value cut off = 0.05. Whiskers from minimum to maximum, median abundances and interquartile ranges have been indicated in the plots.

**Figure 4.**
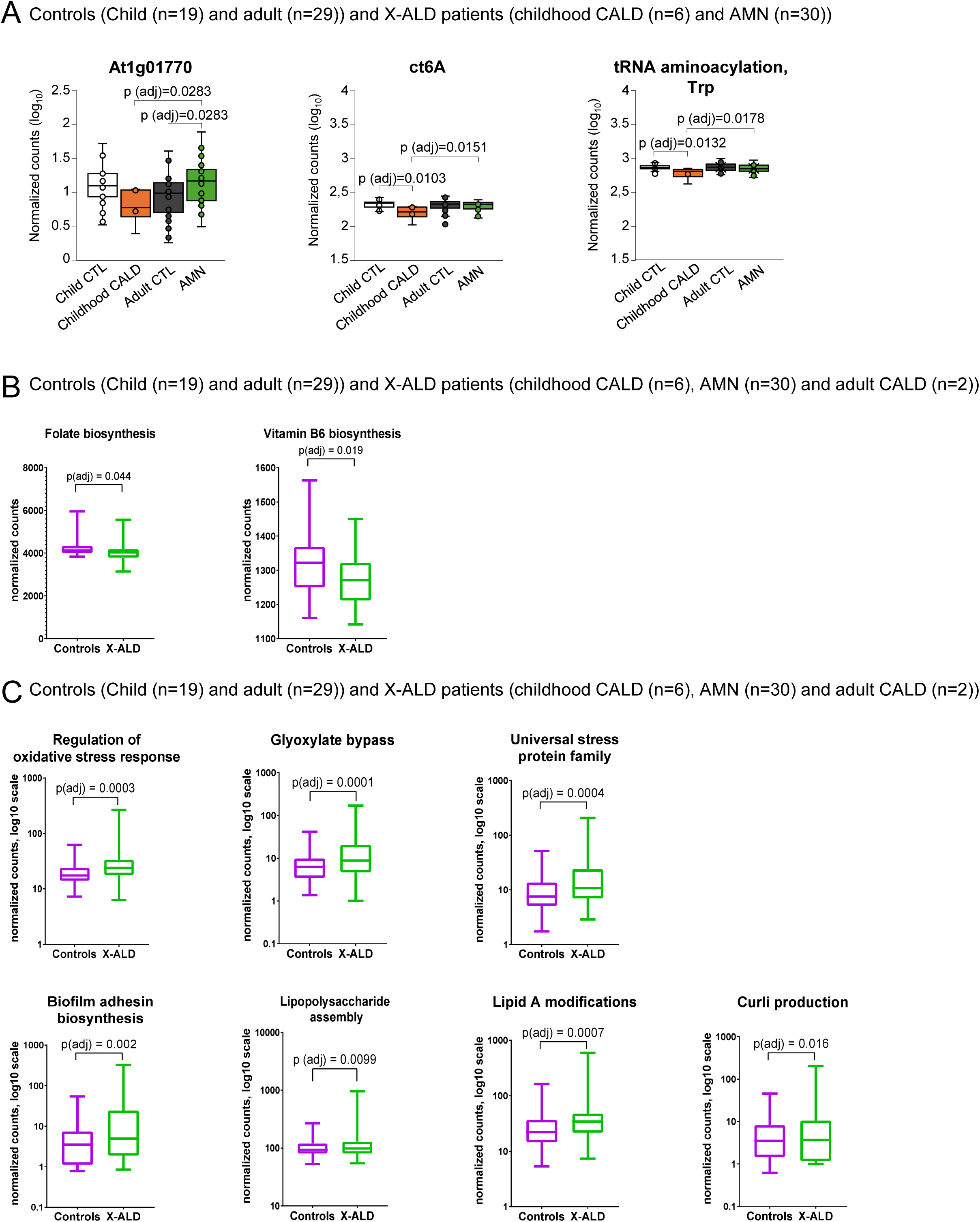
Microbiota functional features of environment adaptation in X-ALD patients. **(A)** Box plots showing the differential abundances (normalised read counts on log10 scale) of functions involved in gut microbiota environmental response in childhood CALD patients compared to AMN (childhood CALD (n=6), AMN (n=30) respect to healthy patients (n= 19 child and n=29 adult)). p-values were calculated with 2-way ANOVA followed by Wilcoxon test. (B) Box plots showing the differential abundances (normalised read counts) of functions implicated in vitamin B6 and folate biosynthesis in the gut microbiota of X-ALD patients compared to controls. Negative binomial models were used for differential abundance testing of subsystem level 3 (function) features. p-values were calculated with LRT test with an adjusted p-value cut off = 0.05. Whiskers from minimum to maximum, median abundances and interquartile ranges have been indicated in the plots. **(C)** Box plots showing the differential abundances (normalised read counts on log10 scale) of functions involved in gut microbiota environmental response in X-ALD patients (n=38) compared to controls (n=48). Negative binomial models were used for differential abundance testing of subsystem level 3 (function) features. p-values were calculated with LRT test with an adjusted p-value cut off = 0.05. Whiskers from minimum to maximum, median abundances and interquartile ranges have been indicated in the plots.

**Table 1.**
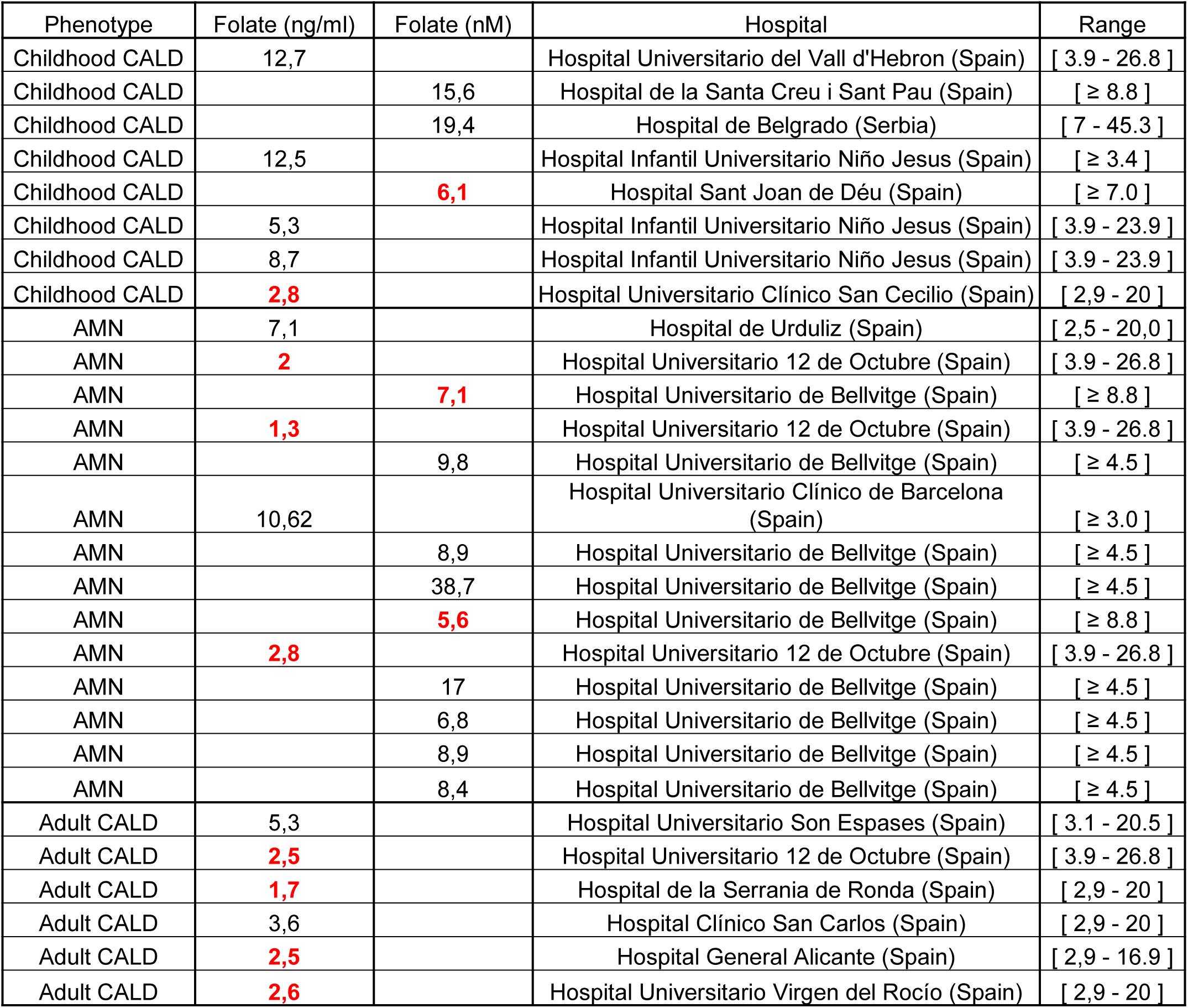
Prevalence of folate deficiency across a multicenter cohort. Serum folate concentrations (ng/mL and nM) were obtained from routine clinical analyses performed at participating hospitals in X-ALD patients (childhood CALD (n=8), AMN (n=14), and adult CALD patients (n=6)). Reference ranges correspond to hospital-specific laboratory cutoffs. Folate deficiency was defined according to each center’s lower reference limit. Overall, 11 of 28 patients (39.3%) were classified as folate deficient. The observed prevalence was significantly higher than expected in the general population (exact one-sample binomial test, p < 0.0001).

| Phenotype | Folate (ng/ml) | Folate (nM) | Hospital | Range |
| --- | --- | --- | --- | --- |
| Childhood CALD | 12,7 |  | Hospital Universitario del Vall d'Hebron (Spain) | [ 3.9 - 26.8 ] |
| Childhood CALD |  | 15,6 | Hospital de la Santa Creu i Sant Pau (Spain) | [ ≥ 8.8 ] |
| Childhood CALD |  | 19,4 | Hospital de Belgrado (Serbia) | [ 7 - 45.3 ] |
| Childhood CALD | 12,5 |  | Hospital Infantil Universitario Niño Jesus (Spain) | [ ≥ 3.4 ] |
| Childhood CALD |  | **6,1** | Hospital Sant Joan de Déu (Spain) | [ ≥ 7.0 ] |
| Childhood CALD | 5,3 |  | Hospital Infantil Universitario Niño Jesus (Spain) | [ 3.9 - 23.9 ] |
| Childhood CALD | 8,7 |  | Hospital Infantil Universitario Niño Jesus (Spain) | [ 3.9 - 23.9 ] |
| Childhood CALD | **2,8** |  | Hospital Universitario Clínico San Cecilio (Spain) | [ 2,9 - 20 ] |
| AMN | 7,1 |  | Hospital de Urduliz (Spain) | [ 2,5 - 20,0 ] |
| AMN | **2** |  | Hospital Universitario 12 de Octubre (Spain) | [ 3.9 - 26.8 ] |
| AMN |  | **7,1** | Hospital Universitario de Bellvitge (Spain) | [ ≥ 8.8 ] |
| AMN | **1,3** |  | Hospital Universitario 12 de Octubre (Spain) | [ 3.9 - 26.8 ] |
| AMN |  | 9,8 | Hospital Universitario de Bellvitge (Spain) | [ ≥ 4.5 ] |
| AMN | 10,62 |  | Hospital Universitario Clínico de Barcelona (Spain) | [ ≥ 3.0 ] |
| AMN |  | 8,9 | Hospital Universitario de Bellvitge (Spain) | [ ≥ 4.5 ] |
| AMN |  | 38,7 | Hospital Universitario de Bellvitge (Spain) | [ ≥ 4.5 ] |
| AMN |  | **5,6** | Hospital Universitario de Bellvitge (Spain) | [ ≥ 8.8 ] |
| AMN | **2,8** |  | Hospital Universitario 12 de Octubre (Spain) | [ 3.9 - 26.8 ] |
| AMN |  | 17 | Hospital Universitario de Bellvitge (Spain) | [ ≥ 4.5 ] |
| AMN |  | 6,8 | Hospital Universitario de Bellvitge (Spain) | [ ≥ 4.5 ] |
| AMN |  | 8,9 | Hospital Universitario de Bellvitge (Spain) | [ ≥ 4.5 ] |
| AMN |  | 8,4 | Hospital Universitario de Bellvitge (Spain) | [ ≥ 4.5 ] |
| Adult CALD | 5,3 |  | Hospital Universitario Son Espases (Spain) | [ 3.1 - 20.5 ] |
| Adult CALD | **2,5** |  | Hospital Universitario 12 de Octubre (Spain) | [ 3.9 - 26.8 ] |
| Adult CALD | **1,7** |  | Hospital de la Serrania de Ronda (Spain) | [ 2,9 - 20 ] |
| Adult CALD | 3,6 |  | Hospital Clínico San Carlos (Spain) | [ 2,9 - 20 ] |
| Adult CALD | **2,5** |  | Hospital General Alicante (Spain) | [ 2,9 - 16.9 ] |
| Adult CALD | **2,6** |  | Hospital Universitario Virgen del Rocío (Spain) | [ 2,9 - 20 ] |

### Methionine and BCAAs: overlooked players in the pathogenesis of X-ALD?

Our discovery of the identification of a gut microbial signature indicating disrupted amino acid metabolism, reinforces the hypothesis that the gut microbiota may contribute to X-ALD pathophysiology. To further investigate this link, we assessed the plasma amino acid profiles of patients (Fig. 5 and Fig. S2).

**Figure 5.**
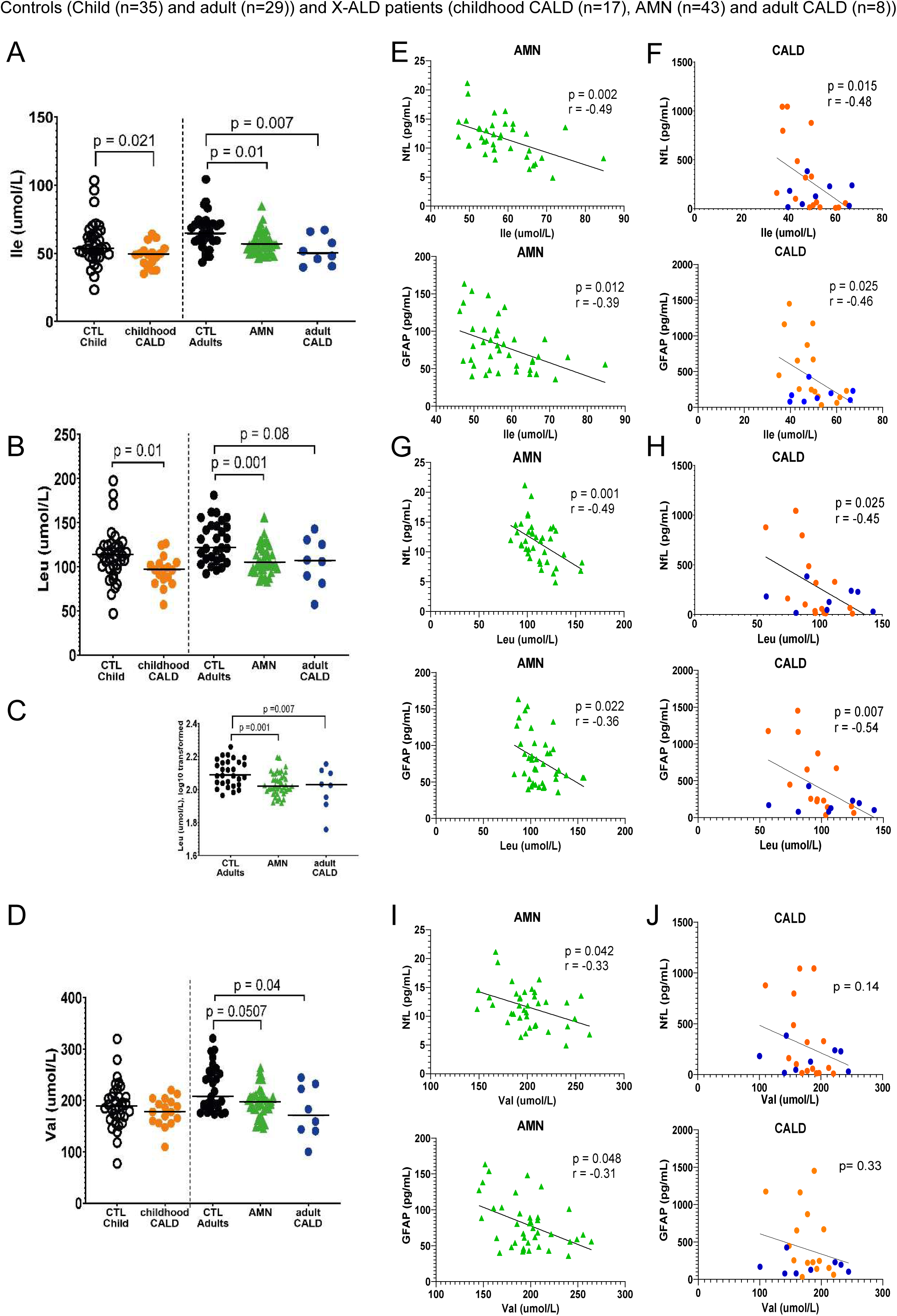
X-ALD BCAA signature in plasma. Dot plots showing the different concentrations (umol/L) of the amino acids isoleucine (Ile) (A), leucine (Leu) (B,C) and valine (Val) (D) (i.e. branched-chain amino acids, BCAA) in X-ALD patients (childhood CALD, n = 17; AMN, n = 43; adult CALD, n = 8) respect to controls (control child n = 35; control adult n = 29). The median of the amino acid levels in each group is indicated by a horizontal line. Outliers were removed with Rout test. An unpaired t-test was used to compare Val in childhood CALD patients with respect to their controls, while the analogous non-parametric Mann-Whitney test was used to compare Ile and Leu levels in childhood CALD patients with respect to their controls. The non-parametric Kruskal-Wallis test was used to compare Ile, Leu and Val levels in adult CALD and AMN patients with respect to their controls. For Leu, a one-way ANOVA was also performed on log transformed levels (B1b). p value threshold was set at 0.05. The scatter plots are showing the correlation between the plasma concentrations of isoleucine (Ile) (E, F), leucine (Leu) (G, H) and valine (Val) (I, J), (umol/L, X axis) and the variables NfL or GFAP (pg/ml, Y axis) in X-ALD patients. p value threshold was set at 0.05. CALD patients (childhood and adult CALD) are represented on the graphs on the right whereas AMN on the left. Orange dots represents childhood CALD patients whereas blue dots represent adult CALD patients. The straight black line represents the line of best fit between the values.

Targeted analysis revealed significantly lower concentrations of essential amino acids in X-ALD patients plasma compared, particularly the branched-chain amino acids (BCAAs) isoleucine (Ile), leucine (Leu), and valine (Val)(Fig. 5A-D). All three BCAAs were significantly lower in adult CALD and/or AMN patients relative to adult controls, while Ile and Leu were also significantly decreased in childhood CALD patients (Fig. 5A-B). Notably, these findings are consistent with the observed reduction in microbial BCAA biosynthesis pathways identified through shotgun metagenomic analysis (Fig. 3). Neurofilament light chain (NfL) and glial fibrillary acidic protein (GFAP) are established biomarkers of neuronal and astrocytic injury, respectively ^50,51^. In X-ALD, both biomarkers are elevated in childhood CALD and correlate with disease severity, whereas in AMN and adult CALD, only NfL is significantly increased and shows an association with disease severity ^37^. We observed an inverse correlation between plasma levels of Leu and Ile and those of NfL and GFAP in both cerebral and AMN forms of the disease (Fig. 5E-H). For Val, this inverse correlation was only detected in AMN patients (Fig. 5I-J). Receiver operating characteristic (ROC) analysis demonstrated that Ile and Leu could effectively distinguish patients with childhood and adult CALD, and AMN from their respective controls. In contrast, Val could differentiate adult CALD and AMN patients but failed to distinguish childhood CALD cases. Among the BCAAs, Ile showed the highest discriminative performance, particularly for adult CALD patients (AUC = 79.74%), whereas Val showed the lowest accuracy in separating AMN patients from controls (AUC = 65.84%) (see Supplementary Material, Fig. S3).

Methionine (Met) levels were also reduced in patients with cerebral forms of the disease (childhood and adult CALD) compared to controls (Fig. 6A). Importantly, we observed an inverse correlation between plasma Met and NfL levels in CALD forms, but not in AMN patients (Fig. 6B-C). ROC analysis indicated that Met effectively discriminated CALD patients from their controls (AUC = 70.41%), but failed to differentiate AMN patients from their own control group (see Supplementary Material, Fig. S3).

**Figure 6.**
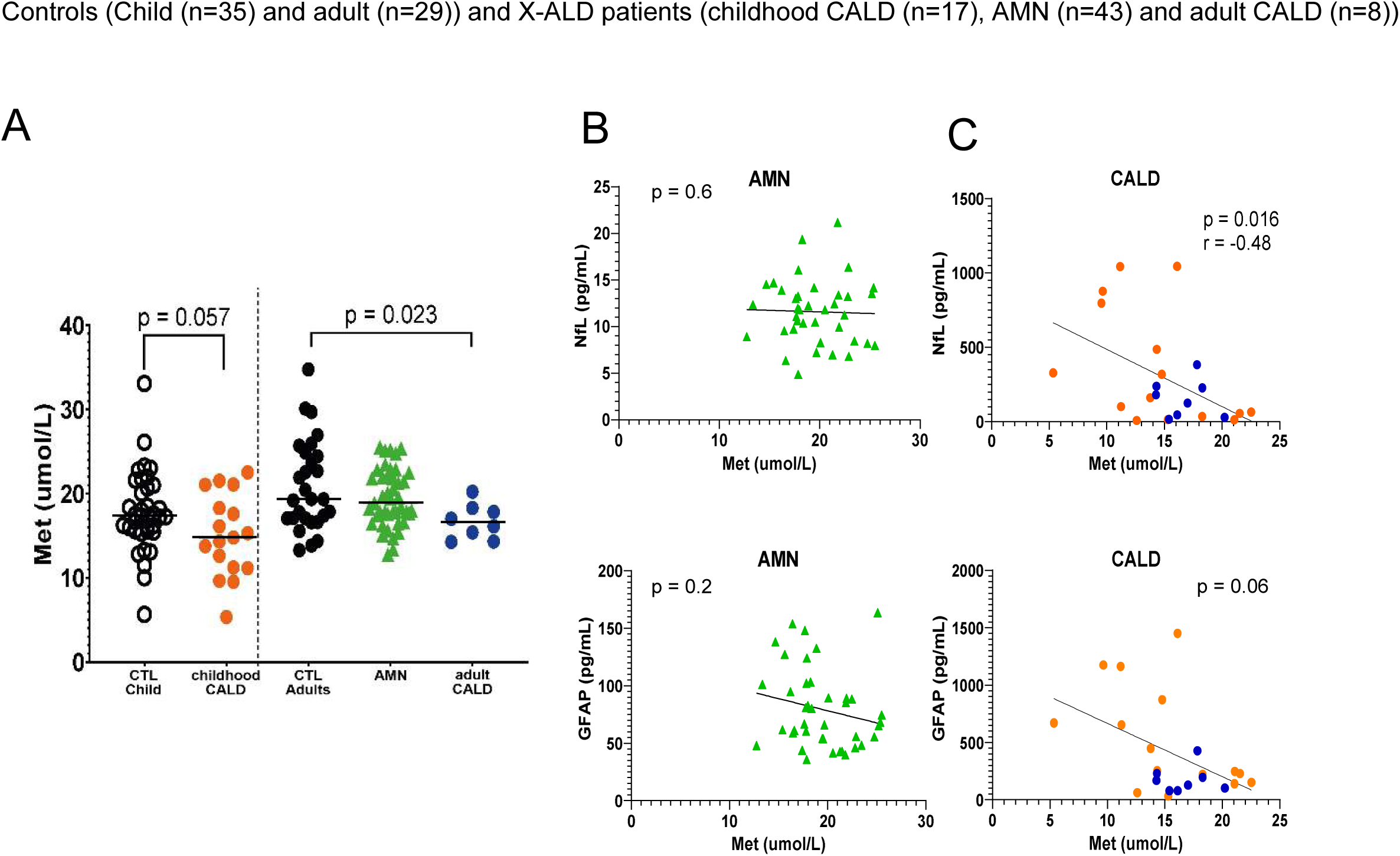
X-ALD methionine signature in plasma. Dot plots showing the different concentrations (umol/L) of the amino acid methionine (Met) in X-ALD patients (childhood CALD, n = 17; AMN, n = 43; adult CALD, n = 8) respect to controls (control child n = 35; control adult n = 29) (A). The median of the amino acid levels in each group is indicated by a horizontal line. Outliers were removed with Rout test. An unpaired t-test was used to compare Met levels in childhood CALD patients with respect to their controls. One-way ANOVA was used to compare Met levels in adult CALD and AMN patients with respect to their controls. p value threshold was set at 0.05. The scatter plots are showing the correlation between the plasma concentrations of methionine (Met) (umol/L, X axis) and the variables NfL or GFAP (pg/ml, Y axis) in X-ALD patients (B, C). p value threshold was set at 0.05. CALD patients (childhood and adult CALD) are represented on the graphs on the right whereas AMN on the left. Orange dots represents childhood CALD patients whereas blue dots represent adult CALD patients. The straight black line represents the line of best fit between the values.

## DISCUSSION

In this study, we identified for the first time two bacterial species that are differentially abundant between AMN and childhood CALD patients. Among these, *H. apodemus*, a pathobiont with reported pro-inflammatory activity mediated through ILC3-associated pathways, was significantly reduced in AMN patients. Although this pattern has not been previously described in other neurodegenerative diseases, the decreased abundance of *H. apodemus* in AMN may reflect a protective microbiota profile associated with the absence of cerebral demyelination in this patient group. This observation supports the potential involvement of a microbiota–gut–brain axis in modulating neuroinflammatory processes in X-ALD, as previously described in other neurodegenerative and neuroinflammatory disorders ^23–28^. On the other hand, the increased abundance of *B. cellulosilyticus*, a metabolically beneficial commensal bacterium, in AMN patients may contribute to protection against CALD-associated demyelination through enhanced riboflavin biosynthesis.

Gut bacteria are not only capable of synthesizing and degrading amino acids for their own metabolic needs but also of producing de novo amino acids, including essential amino acids for the host, thereby establishing a symbiotic relationship with the human body ^52,53^, although the contribution of such biosynthesis to the total amino acid pool in the human organisms is not yet established. In our X-ALD patient cohort, we identified a significant reduction in bacterial functions involved in the biosynthesis of branched-chain amino acids (BCAAs) and methionine (Met) (Fig. 3). It is intriguing that we observed lower plasma levels of these amino acids in X-ALD patients (Fig. 5-6), with BCAAs concentrations inversely correlating with plasma levels of NfL and GFAP, two established biomarkers of neuronal and astrocytic damage, respectively ^50,51^. Elevated NfL and GFAP levels have been previously linked to inflammatory activity and disease progression in cerebral forms of X-ALD and to spinal cord degeneration in AMN patients ^54–56^. Interestingly, reduced plasma methionine levels were specifically observed in patients with cerebral forms of the disease (childhood and adult CALD) and correlated inversely with NfL levels in these subgroups (Fig. 6A, C). BCAAs constitute approximately 35% of essential amino acids in mammals and play crucial roles beyond protein synthesis. Through the branched-chain α-keto acid dehydrogenase (BCKDH) complex in the inner mitochondrial membrane, BCAAs are catabolized and contribute carbon skeletons to the tricarboxylic acid (TCA) cycle. Valine is glucogenic, leucine is ketogenic, and isoleucine has mixed metabolic properties ^57^. BCAAs are also central to inter-organ nitrogen exchange and contribute to the astrocyte-neuron glutamine/glutamate cycle, also referred to as the “leucine-glutamate cycle,” which helps buffer brain glutamate levels and prevent excitotoxicity ^58^. Moreover, enhanced catabolism of BCAAs has been shown to promote M2-like macrophage polarization ^59^, indicating a potential immunomodulatory pathway that could counteract neuroinflammatory processes in X-ALD. Indeed, ABCD1 deficiency impairs macrophage plasticity, hindering the full establishment of anti-inflammatory M2 phenotypes ^60^. This dysfunction may contribute to the severe and rapidly progressive demyelination observed in CALD, a process that sometimes undergoes spontaneous arrest.

In any case, the levels of Val, Ile, and Leu in the blood are efficiently regulated by other factors beyond their production by the microbiota, such as dietary intake, fasting duration, and mitochondrial catabolism. Furthermore, although significant differences in plasma amino acid concentrations were observed between groups, individual values showed substantial overlap. Thus, the correlation observed among amino acids, NfL and GFAP is likely to be more indicative of microbiota dysregulation than the blood concentration of branched chain amino acids. Importantly, because valine and isoleucine catabolism contributes propionyl-CoA and succinyl-CoA to the TCA cycle, such dysregulation may indirectly signal impaired anaplerotic support of the TCA cycle, a vulnerability often implicated in neurodegenerative conditions ^61^.Methionine, a sulfur-containing essential amino acid obtained from the diet or produced by gut microbiota, serves as the initiator of protein synthesis in both prokaryotic and eukaryotic systems. It also acts as a surface antioxidant in proteins. Methionine is metabolized to S-adenosylmethionine (SAM), a key methyl donor required for numerous biosynthetic pathways, including the production of phosphatidylcholine, a critical component of cell membranes ^62,63^. Deficiencies in SAM have been associated with demyelination in certain inborn errors of methylation ^63^. Moreover, prior studies have identified polymorphisms in genes encoding methionine synthase (MTR), cystathionine β-synthase (CBS), and the methionine transporter transcobalamin 2, all involved in methionine metabolism, as possible contributors to phenotypic variability in X-ALD, predicting that higher methionine would protect from inflammatory disease ^64–66^. The inverse correlation we observed between plasma levels of Met and NfL in CALD patients further supports the potential neuroprotective role of this amino acid. Our findings suggest that impaired *de novo* synthesis of amino acids by the gut microbiota may contribute to systemic deficiencies in BCAAs and Met, with possible deleterious consequences for the nervous system through metabolic and immunometabolic mechanisms. Finally, one of the most important determinants of methionine production is folate. As the microbiota may influence folate status in the body, this may establish a relationship between low folate production in patients and significantly reduced methionine levels.

Beyond metabolic alterations, our metagenomic analysis revealed an increased abundance of bacterial pathways involved in oxidative stress response, glyoxylate bypass, universal stress protein expression, and biofilm adhesin production (Fig. 4C), indicating microbial adaptation to a redox-stressed environment. Notably, enrichment of pathways related to oxidative stress responses and biofilm adhesins has also been reported in other neurodegenerative disorders ^67–69^. Thus, we posit that a persistent redox imbalance of mitochondrial origin ^5,7,70,71^ likely imposes a chronic oxidative pressure on both host cells and associated microbial communities in X-ALD. Consistent with this, we observed a marked enrichment of pathways involved in oxidative stress regulation, including key redox-responsive regulators and detoxifying enzymes, alongside a significant increase in universal stress protein (USP) family gene expression. The upregulation of these stress adaptation modules may represent a survival strategy, enabling microbes to withstand elevated reactive oxygen species and other stressors in a compromised host environment. Importantly, the coexistence of proteasome dysfunction and impaired autophagy in X-ALD ^72,73^ could exacerbate the oxidative burden and ER stress that we demonstrated in this disease ^74^, by impairing the clearance of damaged proteins and organelles, thereby perpetuating a cycle of cellular dysfunction. This altered proteostatic landscape may, in turn, select for microbial populations with enhanced stress resistance capacities, as reflected by the activation of oxidative defense and USP pathways. Together, these findings suggest that disruption of redox and proteostasis networks in the host not only contributes to disease progression but also shapes the functional potential of the microbiota, potentially influencing disease outcomes and response to therapy. These results suggest a state of redox imbalance in the gastrointestinal tract of X-ALD patients ^75–77^. Consistent with this interpretation, studies using *Drosophila melanogaster* have demonstrated that peroxisomal dysfunction can lead to redox imbalance and lipotoxicity, which in turn alters gut microbiota composition ^78^. Redox imbalance can also disrupt intestinal epithelial cell function, leading to barrier dysfunction and increased translocation of microbial products into systemic circulation ^79,80^. This compromised gut barrier could exacerbate systemic inflammation and immune activation. Notably, our results show increased abundance of lipopolysaccharide (LPS) biosynthesis and lipid A modification pathways (Fig. 4B), which could represent bacterial responses to oxidative and immune stress. LPS is a key component of the outer membrane of Gram-negative bacteria and acts as a pathogen-associated molecular pattern (PAMP) that can activate the host’s innate immune response through Toll-like receptors (TLRs), particularly TLR4 and TLR2 ^81–83^. Modifications in lipid A structure, especially in the number and length of its acyl chains, can modulate the immunogenic potential of LPS, shifting it between pro-inflammatory, weakly stimulatory, or even antagonistic responses ^81–83^. The potential role of LPS in X-ALD pathophysiology is further supported by our previous observation of elevated TLR2 and CD14 expression in the non-affected white matter of adult CALD patients ^84^.

We also detected increased abundance of *Escherichia coli* (Fig. 2B) and enhanced Curli protein production (Fig. 4B) in X-ALD patients. *E. coli* thrives in inflammatory environments and produces Curli proteins. These proteins can activate the NLRP3 inflammasome and TLRs 1 and 2, further promoting inflammation ^85–87^. Additionally, we identified increased abundance of *Streptococcus mutans* (Fig. 2B), a species known to produce amyloid-like proteins ^88,89^. Recent research has implicated bacterial amyloids such as Curli in the pathogenesis of neurodegenerative diseases, including Parkinson’s disease and amyotrophic lateral sclerosis (ALS) ^90,91^. These parallels warrant further investigation in the context of X-ALD.

This study advances a novel hypothesis linking gut microbiota dysbiosis to the metabolic alterations observed in X-ALD. Using a multi-omics approach in human subjects, we identified disruptions in microbial amino acid biosynthesis, systemic amino acid profiles in particular Methionine, redox stress responses, and immune-related bacterial pathways. Notably, two bacterial species were differentially abundant in AMN compared with childhood cerebral CALD and may reflect protective features. Specifically, *H. apodemus*, a pro-inflammatory species, was reduced in AMN, potentially limiting its deleterious immunomodulatory effects and thereby lowering the likelihood of triggering inflammatory cascades that can contribute to demyelination. In parallel, the anti-inflammatory species *B. cellulosilyticus*, reported to promote regulatory immune responses and to produce riboflavin, was increased in AMN patients. Overall, these findings support the concept that microbial dysregulation may contribute to neurodegeneration and immune imbalance in X-ALD, and they motivate mechanistic and interventional studies to evaluate causality and therapeutic potential. While further studies are needed to elucidate causality and mechanisms, our results point toward exploring the potential of a multi-pronged therapeutic strategy including anti-inflammatory microbiota strains. Such an approach could aim to restore amino acid homeostasis, support anaplerotic replenishment of the TCA cycle, and target gut microbiota composition and intestinal inflammation. By addressing both metabolic and immunological imbalances, this strategy may offer synergistic and disease-modifying effects in the treatment of X-ALD.

## Supporting information

ES1_taxa_general_sequential_model

ES2_functions_general_sequential_model

## Data Availability

The shotgun metagenomic sequencing data have been deposited in the European Nucleotide Archive under accession number PRJEB113847. All data produced in the present study are available upon reasonable request to the authors

https://www.ebi.ac.uk/ena/browser/home

## ACKNOWLEDGEMENTS

We are indebted to patients and families and the ELA-España for their collaboration. We thank the CERCA Program/Generalitat de Catalunya for institutional support. Part of the work presented in this manuscript was previously included in the doctoral thesis of the first author, Lorenzo Torreni (Lorenzo Torreni. Elucidating variable disease severity of X-linked adrenoleukodystrophy through multiomics and metagenomics of gut microbiome. University of Barcelona. Doctoral thesis, 19/11/2024).

## AUTHOR CONTRIBUTIONS

S.F. and A.P. conceived the study, developed the research plan, and supervised the project. L.T., A.P., and S.F. wrote the manuscript. V.V-S., A.R.-P., F.M., S.A.-A., C.C., V.C., and W.K. provided patient samples and collected relevant clinical data. An.A. and R.A. performed amino acid quantification. L.T., A.S., M.R., Ay.A., C.G., and S.M. conducted data analysis. S.F. and A.P. acquired funding. All authors critically reviewed the manuscript and approved the final version for submission.

## DATA AVAILABILITY STATEMENT

The shotgun metagenomic sequencing data have been deposited in the European Nucleotide Archive under accession number PRJEB113847.

## DECLARATION OF GENERATIVE AI AND AI ASSISTED TECHNOLOGIES IN THE WRITING PROCESS

While preparing this work, the author(s) used the AI-powered language model ChatGPT (GPT-4o) to improve readability and language clarity. After using these tools, the authors reviewed and edited the content as needed and take full responsibility for the publication’s content.

## SUPPLEMENTAL FIGURES

**Tables ES1, ES2:** tables containing the list of taxa and functions, respectively, found significantly different in X-ALD patients compared to controls.

**Figure S1.**
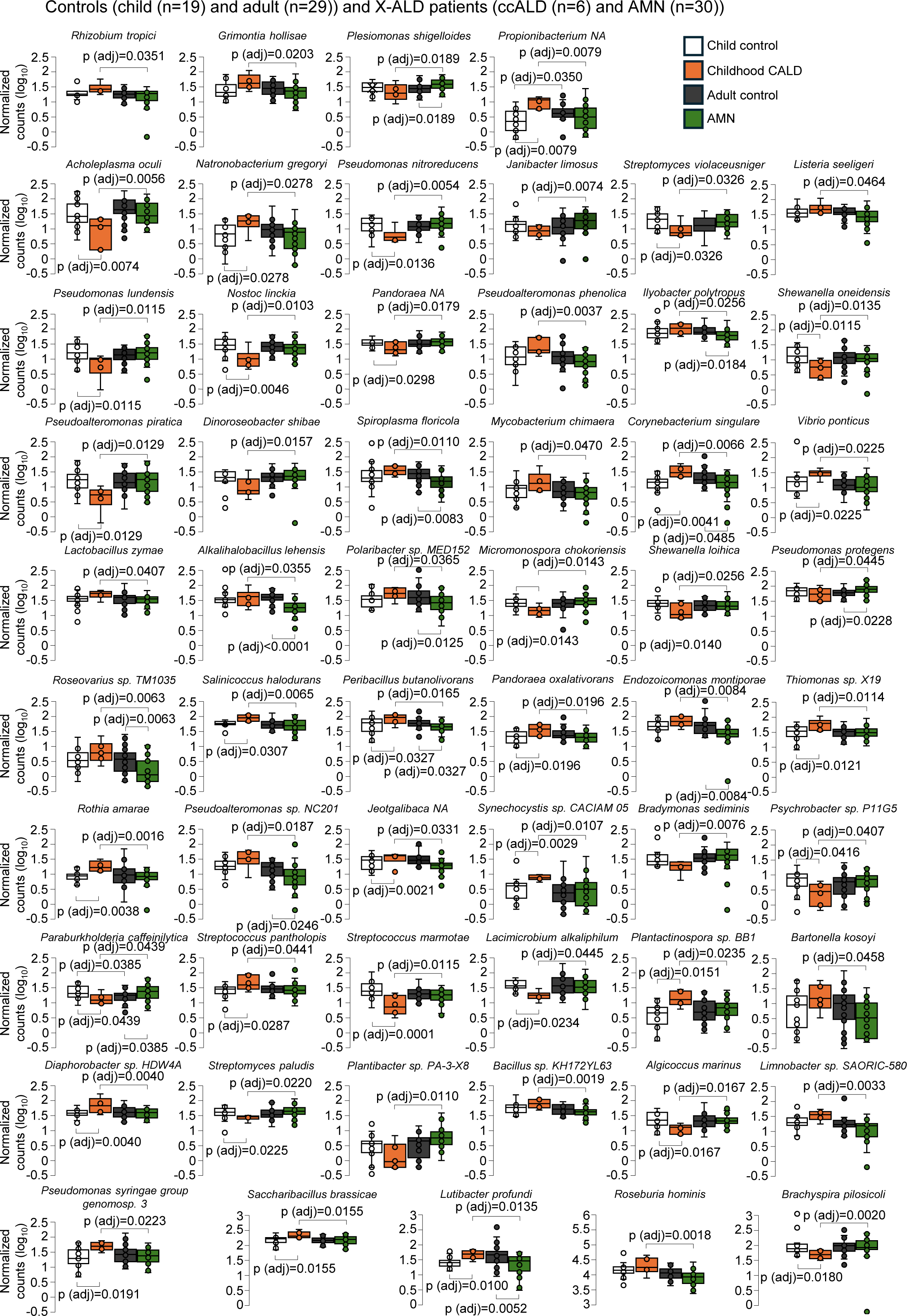
The 58 species level altered between in childhood CALD and AMN microbiota. Box plots showing the differential abundances (normalised read counts on log10 scale) of several species in childhood CALD (n=6), AMN (n=30) respect to healthy patients (n= 19 child and n=29 adult). Whiskers from minimum to maximum, median abundances and interquartile ranges have been indicated in the plots. We looked for differences between among groups, and also for difference in disease status after controlling for age differences. Statistical significance was assessed using 2-way ANOVA or the non-parametric Scheirer–Ray–Hare test (for non-normal data), followed by Wilcoxon rank-sum post-hoc comparisons with Benjamini–Hochberg FDR correction.

**Figure S2.**
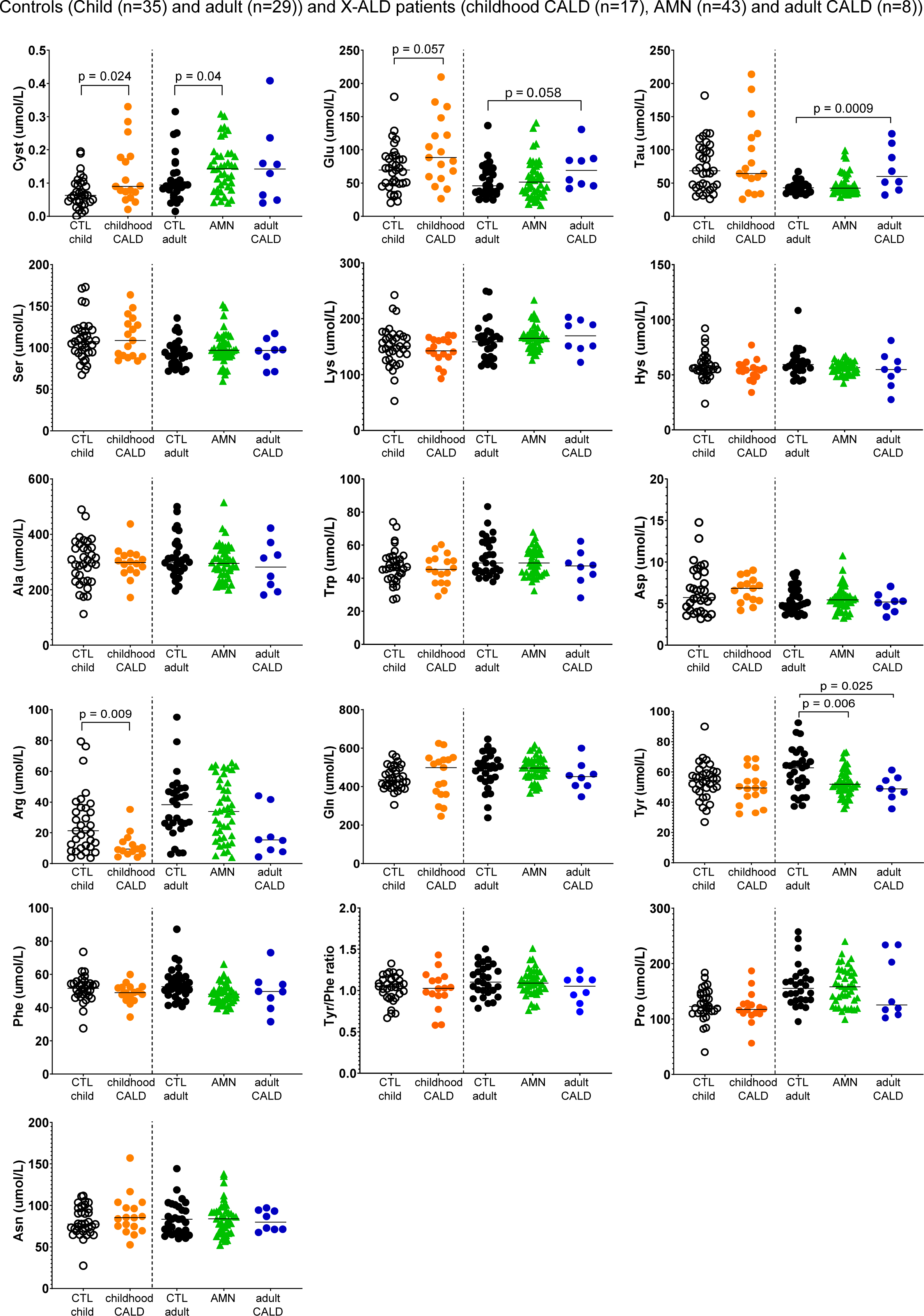
Dot plots showing the concentrations (umol/L) of the other amino acids measured in X-ALD patients (childhood and adult CALD and AMN) and controls. Outliers were removed with Rout test. The p value indicated in the cystine (Cyst), glutamate (Glu) and taurine (Tau) graphs were calculated performing an unpaired t test or the corresponding non-parametric Mann-Whitney test, depending on the distribution of the values. The one-way ANOVA test was used to compare Tyr levels in adult CALD and AMN patients with respect to their controls. An unpaired t-test was used to compare Cys levels in childhood CALD patients with respect to their controls. For Cys levels in the adult groups, the one-way ANOVA test yielded a borderline significant p-value (p = 0.059); therefore, the p-value shown in the graph reflects the result of an unpaired t-test between the adult control and AMN groups. p value threshold was set at 0.05. The median of the amino acid levels in each group is indicated by a horizontal line.

**Figure S3.**
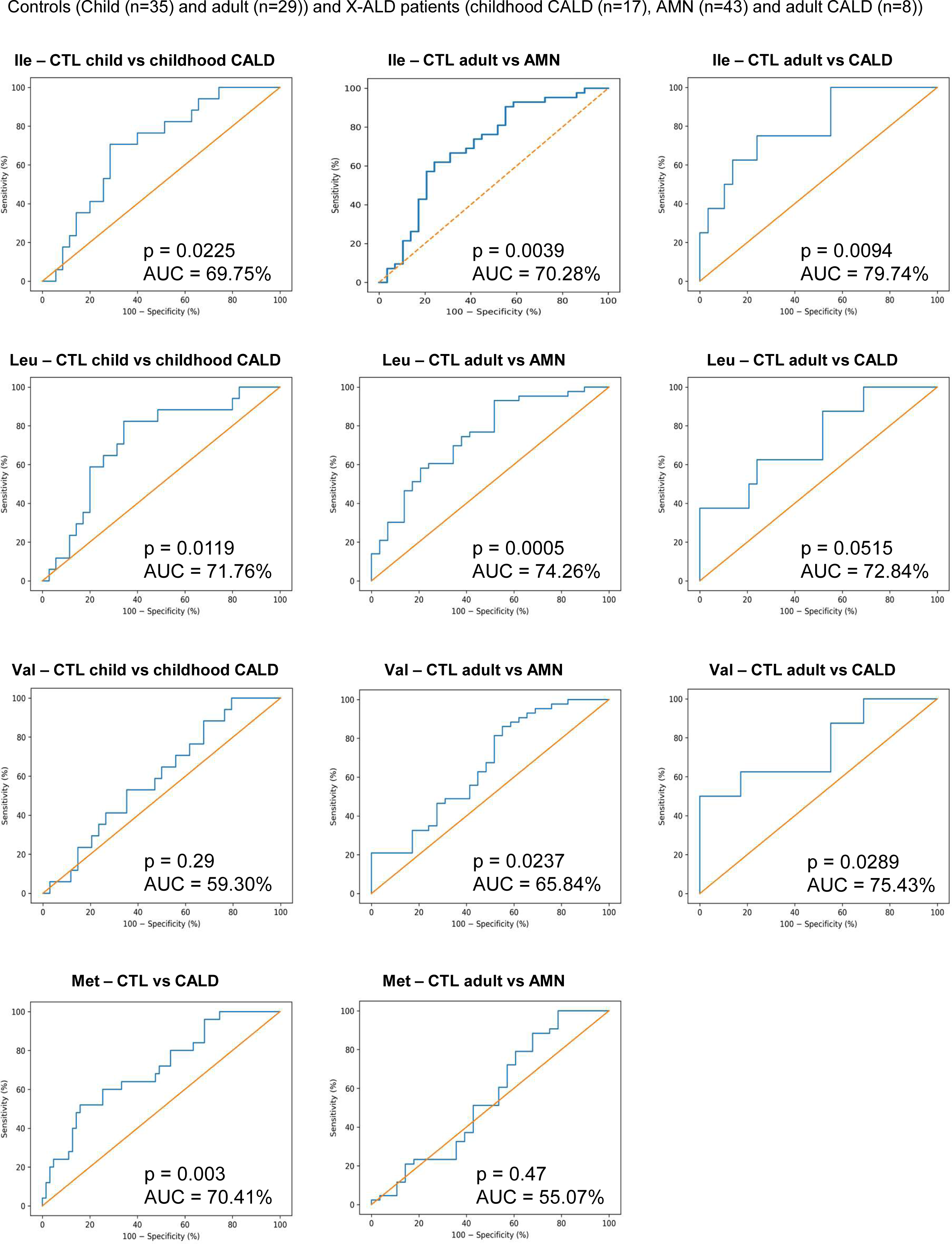
ROC curves with the AUC (in %) comparing each X-ALD phenotype with its age-matched controls for testing the diagnostic accuracy for the plasma levels of the amino acids isoleucine (Ile), leucine (Leu), valine (Val) and methionine (Met). The confidence interval was set at 95% and the p-value threshold was set at 0.05.

