## Supplementary material for "Gut microbiota dysbiosis in adrenoleukodystrophy: a new piece in the puzzle?": ES1_taxa_general_sequential_model

| | baseMean | log2FoldChan | lfcSE | stat | pvalue | padj | Genus | Species |
|---|---|---|---|---|---|---|---|---|
| 1004901 | 19,04847706 | 0,518789768 | 0,146866802 | 12,48339371 | 0,000410586 | 0,027297381 | Gordonia | Gordonia iterans |
| 1028752 | 20,50484882 | -0,6771453 | 0,189317192 | 12,8481331 | 0,000337816 | 0,025409835 | Shewanella | Shewanella aestuarii |
| 1032623 | 13,47562914 | -1,18506862 | 0,276184909 | 17,71785864 | 2,56E-05 | 0,006535581 | Otariodibacter | Otariodibacter oris |
| 1055487 | 9,835454317 | -1,11342271 | 0,31291832 | 12,27714301 | 0,00045854 | 0,027761739 | Methylotenera | Methylotenera versatilis |
| 106634 | 28,0982046 | 0,656620209 | 0,174831707 | 13,65346185 | 0,000219836 | 0,019941876 | Thioalkalivibrio | Thioalkalivibrio versutus |
| 107709 | 58,86986891 | 0,412988575 | 0,117920851 | 11,97952443 | 0,000537883 | 0,028688728 | Opitutus | Opitutus terrae |
| 108980 | 12,23986515 | -1,07927594 | 0,27855269 | 15,05515433 | 0,000104415 | 0,014056218 | Acinetobacter | Acinetobacter ursingii |
| 112234 | 45,21369571 | -0,74902408 | 0,207358046 | 13,28935315 | 0,000266918 | 0,022260127 | Chryseobacte | Chryseobacterium joostei |
| 1129704 | 22,97616622 | 0,509392323 | 0,143868529 | 12,86306659 | 0,000335131 | 0,025409835 | Kyrpidia | NA |
| 1141221 | 40,53224915 | -0,70924301 | 0,213862443 | 11,21118033 | 0,00081306 | 0,034921856 | Chryseobacte | Chryseobacterium taihuense |
| 1144748 | 17,22225206 | -0,80524789 | 0,228526984 | 12,55509178 | 0,000395128 | 0,027297381 | Kangiella | Kangiella sediminilitoris |
| 1176482 | 34,73017541 | -0,77691273 | 0,234130439 | 10,97827564 | 0,000921862 | 0,036622114 | Sulfurimonas | Sulfurimonas gotlandica |
| 1177 | 28,18695227 | -0,5420428 | 0,172479891 | 9,710482399 | 0,001832199 | 0,049543345 | Nostoc | NA |
| 119981 | 42,09981479 | 0,356819943 | 0,1069208 | 11,2795594 | 0,000783652 | 0,034865386 | Propionibacte | Propionibacterium australiense |
| 1218801 | 40,04636242 | -0,80225158 | 0,20068841 | 16,399094 | 5,13E-05 | 0,010380493 | Kordia | Kordia antarctica |
| 1222016 | 35,95692351 | -0,48709176 | 0,15520758 | 9,876040811 | 0,001674449 | 0,04669 | Caldicellulosi | Caldicellulosiruptor changbaiensis |
| 1249 | 9,551008701 | -0,72602285 | 0,210953776 | 11,98604047 | 0,000536006 | 0,028688728 | Weissella | Weissella paramesenteroides |
| 1250205 | 39,20629951 | -0,69692762 | 0,224312067 | 9,871273619 | 0,001678793 | 0,04669 | Gramella | Gramella sp. MAR_2010_147 |
| 126385 | 19,20361465 | -0,7967925 | 0,244214689 | 10,391107 | 0,001266237 | 0,04166486 | Providencia | Providencia alcalifaciens |
| 1283 | 58,23963458 | -0,5379743 | 0,161468877 | 11,29698801 | 0,000776329 | 0,034865386 | Staphylococcu | Staphylococcus haemolyticus |
| 1301 | 2242,707192 | 0,78129225 | 0,219136327 | 13,06958536 | 0,00030013 | 0,023795408 | Streptococcus | NA |
| 1304 | 1584,175841 | 1,973013984 | 0,476630149 | 16,78534905 | 4,19E-05 | 0,009094983 | Streptococcus | Streptococcus salivarius |
| 1309 | 54,66694371 | 0,663386774 | 0,204405615 | 10,7434251 | 0,001046505 | 0,039107278 | Streptococcus | Streptococcus mutans |
| 1316410 | 7,36173712 | 2,439585642 | 0,580569028 | 16,87304753 | 4,00E-05 | 0,009018271 | Streptococcus | Streptococcus sp. HSISS1 |
| 1316411 | 22,82409178 | 2,230996406 | 0,400594896 | 29,65323432 | 5,17E-08 | 0,000101042 | Streptococcus | Streptococcus sp. HSISS2 |
| 1316412 | 28,24365924 | 2,06048486 | 0,39489528 | 26,52527739 | 2,60E-07 | 0,000305216 | Streptococcus | Streptococcus sp. HSISS3 |
| 1324352 | 90,34424495 | -0,97952721 | 0,250915622 | 15,48460601 | 8,32E-05 | 0,012722189 | Chryseobacte | Chryseobacterium gallinarum |
| 1335 | 103,3214257 | 1,255618724 | 0,299697204 | 18,19765009 | 1,99E-05 | 0,006490348 | Streptococcus | Streptococcus equinus |
| 1335613 | 236,1090532 | 1,16201984 | 0,240915504 | 24,01870845 | 9,54E-07 | 0,000932894 | Gordonibacter | Gordonibacter urolithinfaciens |
| 1336804 | 30,46750014 | -0,6164259 | 0,196687049 | 9,983515929 | 0,001579478 | 0,045649249 | Polaribacter | Polaribacter sp. Hel1_33_78 |
| 1343 | 81,74890991 | 2,77147411 | 0,398899141 | 43,50137323 | 4,24E-11 | 2,49E-07 | Streptococcus | Streptococcus vestibularis |
| 1374 | 12,27668465 | -0,95222172 | 0,270287671 | 12,40010527 | 0,00042931 | 0,02764147 | Planococcus | Planococcus kocurii |
| 1397 | 104,492803 | -0,3835121 | 0,116790366 | 10,96185706 | 0,000930066 | 0,036622114 | Bacillus | Bacillus circulans |
| 142058 | 6,816300555 | 1,407498406 | 0,367963144 | 14,20459165 | 0,00016397 | 0,01738733 | Anaplasma | Anaplasma ovis |
| 1427356 | 38,65378142 | -0,47042204 | 0,149736266 | 9,884803222 | 0,001666494 | 0,04669 | Hyphomicrobi | Hyphomicrobium nitrativorans |
| 1432056 | 13,31216478 | -0,8777033 | 0,276263235 | 10,26379945 | 0,00135666 | 0,042438132 | Mannheimia | Mannheimia sp. USDA-ARS-USMARC-1261 |
| 143223 | 42,87541305 | -0,99837015 | 0,230905243 | 18,46939958 | 1,73E-05 | 0,005958605 | Salegentibacte | Salegentibacter salegens |
| 1433513 | 13,93318626 | 0,867629407 | 0,268875539 | 10,18514535 | 0,001415766 | 0,043261963 | Streptococcus | Streptococcus gwangjuense |
| 1458492 | 4,888888851 | -1,36770507 | 0,40427167 | 11,1789816 | 0,000827292 | 0,034921856 | Myroides | Myroides sp. ZB35 |
| 1472345 | 13,79238776 | -0,93099226 | 0,255655647 | 13,20760939 | 0,000278815 | 0,022408306 | Basilea | Basilea psittacipulmonis |
| 1541959 | 44,17000921 | -0,70810184 | 0,22848172 | 9,682038853 | 0,00186078 | 0,049543345 | Candidatus Izi | Candidatus Izimaplasma sp. HR1 |
| 1550579 | 68,45451166 | -0,93490162 | 0,21942718 | 18,01958483 | 2,19E-05 | 0,006535581 | Mucilaginibac | Mucilaginibacter gotjawali |
| 1564 | 107,3749933 | 0,370339956 | 0,095331601 | 14,85489984 | 0,000116106 | 0,014056218 | Desulfotomac | Desulfotomaculum ruminis |
| 1572645 | 12,33842961 | -0,77848839 | 0,245638191 | 10,07151695 | 0,00150579 | 0,045205918 | Neptunomona | Neptunomonas phycophila |
| 1573719 | 13,16351858 | -0,80576964 | 0,215621229 | 14,24856848 | 0,000160183 | 0,01738733 | Pseudomonas | Pseudomonas sp. S35 |
| 1581011 | 20,66691766 | -0,92542229 | 0,226210282 | 16,54033595 | 4,76E-05 | 0,009979366 | Sulfurospirillu | Sulfurospirillum sp. UCH001 |
| 1601 | 40,60637605 | 0,659674138 | 0,202708187 | 10,00180669 | 0,001563867 | 0,045649249 | Ligilactobacill | Ligilactobacillus agilis |
| 1609758 | 32,12724683 | 0,495422591 | 0,156461604 | 10,39410519 | 0,001264182 | 0,04166486 | Novosphingob | Novosphingobium sp. P6W |
| 1612150 | 5,339716733 | -1,13804721 | 0,327631707 | 11,69122534 | 0,000627955 | 0,030198474 | Mycoplasma | Mycoplasma tullyi |
| 1641 | 45,32382967 | -0,55483592 | 0,139060165 | 15,61010854 | 7,78E-05 | 0,012342481 | Listeria | Listeria grayi |
| 165096 | 24,73862039 | -0,68222066 | 0,181283764 | 14,18188569 | 0,000165961 | 0,01738733 | Weissella | Weissella koreensis |
| 1654 | 60,33731078 | 0,371486562 | 0,110023939 | 11,26828842 | 0,000788424 | 0,034865386 | Actinomyces | NA |
| 1660 | 165,0948721 | 0,880519456 | 0,216978695 | 15,35910677 | 8,89E-05 | 0,012884991 | Schaalia | Schaalia odontolytica |
| 1682113 | 18,34046109 | 0,496057431 | 0,148604639 | 11,04287094 | 0,000890289 | 0,036526746 | Mycobacteriu | Mycobacterium sp. YC-RL4 |
| 1763538 | 46,17975038 | -0,6505129 | 0,140453387 | 20,91029761 | 4,81E-06 | 0,002247573 | Paenibacillus | Paenibacillus crassostreae |
| 1778263 | 11,6171388 | -1,40525639 | 0,415716035 | 11,01438547 | 0,000904075 | 0,036580743 | Candidatus Ho | Candidatus Hoaglandella endobia |
| 1789 | 12,74296533 | 0,701529517 | 0,189539406 | 13,64410081 | 0,000220934 | 0,019941876 | Mycobacteriu | Mycobacterium xenopi |
| 1798225 | 24,88090334 | -0,7694238 | 0,243068314 | 10,05016262 | 0,001523345 | 0,045205918 | Formosa | Formosa sp. Hel1_31_208 |
| 1813019 | 13,50390794 | -0,61454375 | 0,193058584 | 10,24814538 | 0,001368221 | 0,042472762 | Campylobacte | Campylobacter hepaticus |
| 1827580 | 22,16027921 | 0,52107013 | 0,143235519 | 13,24270735 | 0,000273642 | 0,022298054 | Streptomyces | Streptomyces nigra |
| 1839799 | 45,20088528 | 2,155438683 | 0,484597445 | 18,63245919 | 1,58E-05 | 0,005811954 | Streptococcus | Streptococcus sp. FDAARGOS_192 |
| 1843210 | 113,4823322 | -0,4909958 | 0,154985348 | 9,933767365 | 0,001622741 | 0,046349306 | Anaerocolumn | NA |
| 184917 | 38,24106311 | 0,425174713 | 0,127908374 | 10,74602879 | 0,001045033 | 0,039107278 | Desulfovibrio | Desulfovibrio magneticus |
| 185007 | 1506,381246 | -0,63880096 | 0,183020207 | 12,01417776 | 0,000527974 | 0,028681685 | Pseudobutyriv | Pseudobutyrivibrio xylanivorans |
| 185008 | 905,0600151 | -0,53413351 | 0,153980016 | 11,88712234 | 0,000565234 | 0,029609168 | Butyrivibrio | Butyrivibrio hungatei |
| 187101 | 38,68992909 | -0,57448727 | 0,173719093 | 11,07543442 | 0,000874789 | 0,036399918 | Sneathia | Sneathia amnii |
| 1873 | 119,8969322 | 0,316812413 | 0,100047974 | 10,023503 | 0,001545552 | 0,045566613 | Micromonosp | NA |
| 1883416 | 24,14284635 | 0,477598433 | 0,130642366 | 13,45778131 | 0,000243992 | 0,021051497 | Halomonas | Halomonas sp. 1513 |
| 1890302 | 12,73538113 | -0,88732938 | 0,277981617 | 10,32052948 | 0,001315585 | 0,042072712 | Bacillus | Bacillus wiedmannii |
| 1891186 | 13,83999652 | -0,95755066 | 0,267415788 | 12,76423576 | 0,00035331 | 0,026071777 | Vibrio | Vibrio aphrogenes |
| 1898474 | 32,45045844 | -0,5421456 | 0,154553401 | 12,2695536 | 0,000460408 | 0,027761739 | Macrococcus | Macrococcus sp. IME1552 |
| 190323 | 7,797342386 | 0,813827359 | 0,249922549 | 10,85397504 | 0,000985839 | 0,038047249 | Plantibacter | NA |
| 1912064 | 11,8922662 | -0,91340368 | 0,277250247 | 10,9634967 | 0,000929243 | 0,036622114 | Staphylococcu | Staphylococcus sp. MI 10-1553 |
| 1913577 | 37,78799101 | -0,79487516 | 0,258071348 | 9,643428689 | 0,001900302 | 0,049543345 | Gramella | Gramella salexigens |
| 192 | 88,40523609 | 0,307504825 | 0,089561683 | 11,92014704 | 0,000555301 | 0,029350908 | Azospirillum | Azospirillum brasilense |
| 1920749 | 4,191638185 | -1,31436667 | 0,33867748 | 14,93000917 | 0,000111574 | 0,014056218 | Candidatus M | Candidatus Mancarchaeum acidiphilum |
| 1960 | 28,73803557 | 0,550679848 | 0,161494128 | 11,17882194 | 0,000827363 | 0,034921856 | Streptomyces | Streptomyces vinaceus |
| 1973486 | 47,61366863 | -0,50518059 | 0,157175556 | 10,25943671 | 0,001359872 | 0,042438132 | Calothrix | Calothrix parasitica |
| 1978566 | 19,86194165 | 0,520503525 | 0,139777116 | 14,06254252 | 0,000176831 | 0,01820114 | Cryobacterium | Cryobacterium sp. LW097 |
| 202747 | 21,57082582 | -0,56587974 | 0,170711587 | 11,05088572 | 0,000886448 | 0,036526746 | Sulfurimonas | Sulfurimonas autotrophica |
| 202950 | 17,68416719 | -0,72244879 | 0,226675798 | 10,37510116 | 0,001277261 | 0,04166486 | Acinetobacter | Acinetobacter baylyi |
| 2041044 | 630,0661521 | 0,422812841 | 0,13074359 | 10,37362804 | 0,001278281 | 0,04166486 | Eubacterium | Eubacterium maltosivorans |
| 2049305 | 42,23882772 | -0,73375903 | 0,232808216 | 9,687893816 | 0,00185486 | 0,049543345 | Changchengzh | Changchengzhania lutea |
| 2066049 | 5,498900286 | 1,190780184 | 0,316050694 | 14,72323855 | 0,000124502 | 0,014056218 | Citrobacter | Citrobacter freundii complex sp. CFNIH2 |
| 2094025 | 34,91740617 | -0,81502792 | 0,208797458 | 15,61581116 | 7,76E-05 | 0,012342481 | Aureitalea | Aureitalea sp. RR4-38 |

| | | | | | | | | |
|---|---|---|---|---|---|---|---|---|
| 2126321 | 36,28334085 | 0,423379762 | 0,129154483 | 11,09059468 | 0,000867667 | 0,036361426 | Nissabacter | Nissabacter sp. SGAir0207 |
| 2153354 | 16,42392766 | 0,664856809 | 0,206389552 | 10,67223158 | 0,001087558 | 0,039631706 | Klebsiella | Klebsiella huaxiensis |
| 2169540 | 38,84856833 | -0,44928855 | 0,139359002 | 10,30988552 | 0,001323195 | 0,042072712 | Lysinibacillus | Lysinibacillus sp. 2017 |
| 217159 | 118,1009036 | -0,39298557 | 0,116714545 | 11,27748191 | 0,000784529 | 0,034865386 | Clostridium | Clostridium carboxidivorans |
| 2172098 | 43,32452644 | -0,59554839 | 0,185574568 | 10,4720416 | 0,001211947 | 0,041629968 | Flavobacteriu | Flavobacterium pallidum |
| 2183896 | 53,33475502 | -0,74792784 | 0,215171935 | 12,2873735 | 0,000456033 | 0,027761739 | Flavobacteriu | Flavobacterium crocinum |
| 2203204 | 22,79732086 | 0,499639733 | 0,144409671 | 12,38247635 | 0,000433382 | 0,02764147 | Streptomyces | Streptomyces sp. WAC 01438 |
| 2220095 | 17,43260508 | 0,521618223 | 0,165143907 | 10,35807158 | 0,001289098 | 0,041690816 | Cryobacteriun | Cryobacterium soli |
| 2268026 | 59,24539153 | -0,64085195 | 0,205541833 | 9,65062472 | 0,001892872 | 0,049543345 | Runella | Runella sp. SP2 |
| 2269374 | 23,86844355 | -0,59208748 | 0,168220965 | 12,10837232 | 0,000501959 | 0,027782975 | Lactobacillus | Lactobacillus terrae |
| 2287 | 7,567201938 | -1,1282677 | 0,346787743 | 10,3908946 | 0,001266383 | 0,04166486 | Saccharolobu | Saccharolobus solfataricus |
| 229545 | 3,732293089 | -1,59817374 | 0,44847565 | 11,77080224 | 0,00060167 | 0,029963805 | Candidatus Pl | Candidatus Phytoplasma Aster yellows witches'-broom phytoplasma |
| 2341117 | 70,67200643 | -0,70494966 | 0,216266213 | 10,62196177 | 0,001117524 | 0,040067538 | Arachidicoccu | Arachidicoccus soli |
| 2358479 | 31,05534546 | -0,85807076 | 0,26410083 | 10,22201925 | 0,001387738 | 0,0428519 | Tenacibaculur | Tenacibaculum sp. DSM 106434 |
| 237 | 97,35204301 | -0,59322791 | 0,18578718 | 10,35207918 | 0,001293289 | 0,041690816 | Flavobacteriu | NA |
| 237258 | 43,55452544 | -0,68594787 | 0,203095051 | 11,70464819 | 0,000623442 | 0,030198474 | Cloacibacteriu | Cloacibacterium normanense |
| 240495 | 40,19602185 | 0,374382293 | 0,114158698 | 10,84206802 | 0,000992199 | 0,038047249 | Pseudonocarc | Pseudonocardia dioxanivorans |
| 2420306 | 6,325821327 | 0,905060552 | 0,218631777 | 16,97708724 | 3,78E-05 | 0,008878801 | Serratia | Serratia sp. FDAARGOS_506 |
| 242600 | 20,34293917 | -1,25349212 | 0,265822299 | 22,48333185 | 2,12E-06 | 0,001437736 | Ichthyobacteri | Ichthyobacterium seriolicida |
| 2448483 | 3,424508538 | -1,5603823 | 0,444623145 | 12,56263778 | 0,000393536 | 0,027297381 | Serratia | Serratia sp. 3ACOL1 |
| 2479546 | 13,33682699 | -0,88493147 | 0,268802147 | 10,59900349 | 0,001131486 | 0,040067538 | Vibrio | Vibrio sp. HBUAS61001 |
| 250 | 77,91221561 | -0,8307623 | 0,21147666 | 15,83930051 | 6,90E-05 | 0,012342481 | Chryseobacte | Chryseobacterium gleum |
| 2500547 | 46,73103386 | -0,59384858 | 0,189890582 | 9,870878774 | 0,001679153 | 0,04669 | Apibacter | Apibacter raozihei |
| 2507162 | 547,7223073 | 0,304271145 | 0,097484164 | 9,74723344 | 0,001795928 | 0,049468119 | Caproiciprodu | Caproiciproducens sp. NJN-50 |
| 2507538 | 27,57036019 | -0,69328679 | 0,196824686 | 12,14532009 | 0,000492112 | 0,027761739 | Muriicola | Muriicola sp. MMS17-SY002 |
| 2545455 | 64,55571891 | -0,69977128 | 0,215026794 | 10,69356177 | 0,00107509 | 0,039631706 | Arachidicoccu | Arachidicoccus sp. B3-10 |
| 2563897 | 11,71200532 | -0,70226266 | 0,223611185 | 9,996386735 | 0,001568477 | 0,045649249 | Acinetobacter | Acinetobacter sp. 10FS3-1 |
| 2579977 | 10,31296251 | 0,990065378 | 0,277715033 | 11,2637138 | 0,000790369 | 0,034865386 | Brevundimona | Brevundimonas sp. SGAir0440 |
| 2583822 | 8,197227051 | 0,7375128 | 0,203986598 | 12,9402304 | 0,000321595 | 0,024826321 | Plantibacter | Plantibacter sp. M259 |
| 2584944 | 1267,341931 | 1,299061086 | 0,387651391 | 9,688289134 | 0,001854461 | 0,049543345 | Sutterella | Sutterella faecalis |
| 2585135 | 20,56219503 | 0,524839324 | 0,150067642 | 12,31060724 | 0,000450391 | 0,027761739 | Georgenia | Georgenia sp. Z294 |
| 2589076 | 22,92168389 | 0,409477082 | 0,133655447 | 9,645220215 | 0,001898449 | 0,049543345 | Paracoccus | Paracoccus sp. AK26 |
| 2598457 | 19,83600471 | 1,177389899 | 0,344350244 | 10,53684131 | 0,001170182 | 0,040958918 | Streptococcus | Streptococcus sp. KS 6 |
| 2602015 | 42,52714293 | 0,442949786 | 0,113936846 | 15,03288288 | 0,000105654 | 0,014056218 | Hypericibacte | Hypericibacter terrae |
| 2610895 | 5241,88106 | 0,509016601 | 0,142852781 | 13,27211707 | 0,000269383 | 0,022260127 | Flintibacter | Flintibacter sp. KGMB00164 |
| 263 | 31,54486271 | -0,84655036 | 0,181152388 | 22,08575534 | 2,61E-06 | 0,001529746 | Francisella | Francisella tularensis |
| 2651974 | 40,18587202 | 0,450772893 | 0,14085735 | 10,29512605 | 0,00133382 | 0,042072712 | Hydrogenopha | Hydrogenophaga sp. BPS33 |
| 266 | 21,66745595 | 0,473165578 | 0,147031068 | 10,44842433 | 0,001227541 | 0,041629968 | Paracoccus | Paracoccus denitrificans |
| 2662065 | 16,91282527 | 0,54317505 | 0,154705045 | 12,62443396 | 0,000380737 | 0,027297381 | Streptomyces | Streptomyces sp. SYP-A7193 |
| 2662364 | 44,98338736 | -1,04137044 | 0,22274262 | 22,40716871 | 2,21E-06 | 0,001437736 | Sphingobacter | Sphingobacterium sp. dk4302 |
| 2663022 | 47,98765931 | -0,45930202 | 0,137521643 | 11,18606508 | 0,000824139 | 0,034921856 | Gracilibacillus | Gracilibacillus sp. SCU50 |
| 2675218 | 16,32123169 | 0,837222572 | 0,193132389 | 18,96266337 | 1,33E-05 | 0,005213869 | Corynebacteri | Corynebacterium sp. 2019 |
| 2686078 | 31,50481756 | -0,72771751 | 0,224900335 | 10,75326197 | 0,001040957 | 0,039107278 | Winogradskye | Winogradskyella schleiferi |
| 2686366 | 53,97279889 | -1,28015048 | 0,263508576 | 23,53353525 | 1,23E-06 | 0,001028866 | Algibacter | Algibacter sp. L3A6 |
| 2690837 | 191,8802932 | 0,435688255 | 0,11989861 | 13,29444579 | 0,000266194 | 0,022260127 | Eggerthella | Eggerthella sp. HF-1101 |
| 2698682 | 18,69775283 | 0,562988232 | 0,15797181 | 12,16631886 | 0,000486602 | 0,027761739 | Lysobacter | Lysobacter oculi |
| 270 | 41,34379131 | 0,454363007 | 0,129743145 | 12,35093237 | 0,000440766 | 0,027761739 | Thermus | NA |
| 2708020 | 17,01089767 | -0,81298485 | 0,222115462 | 13,5856373 | 0,000227923 | 0,020260949 | Legionella | Legionella sp. TUM19329 |
| 2708338 | 21,00087075 | 0,418422695 | 0,132924229 | 9,984289166 | 0,001578815 | 0,045649249 | Salinibacteriu | Salinibacterium sp. ZJ450 |
| 2719023 | 12,33353082 | 0,729909463 | 0,196651218 | 13,92688656 | 0,00019006 | 0,018721908 | Saccharopolys | Saccharopolyspora sp. ASAGF58 |
| 2733571 | 20,99644564 | 0,597202691 | 0,142524606 | 17,8224341 | 2,43E-05 | 0,006535581 | Trueperella | Trueperella sp. 19M2397 |
| 2748319 | 37,64747387 | -1,03569402 | 0,231579965 | 20,30763281 | 6,59E-06 | 0,002763232 | Flavobacteriu | Flavobacterium sp. I3-2 |
| 2748865 | 16,30970393 | -0,96858295 | 0,288536903 | 11,53152009 | 0,00068426 | 0,03237545 | Bdellovibrio | Bdellovibrio sp. KM01 |
| 2750025 | 11,11689965 | 1,0567734 | 0,252105297 | 15,73892594 | 7,27E-05 | 0,012342481 | Streptomyces | Streptomyces sp. CB00271 |
| 2770636 | 10,60883245 | -0,91682748 | 0,259282832 | 12,19844515 | 0,000478294 | 0,027761739 | Mannheimia | Mannheimia sp. ZY190616 |
| 28025 | 327,7292236 | 1,673985593 | 0,36774126 | 21,65899366 | 3,26E-06 | 0,001737056 | Bifidobacteriu | Bifidobacterium animalis |
| 281120 | 15,20071643 | -0,69472266 | 0,198300758 | 12,25385265 | 0,000464299 | 0,027761739 | Candidatus Ar | Candidatus Amoebophilus asiaticus |
| 28172 | 15,08703581 | -0,77869224 | 0,238988788 | 10,40413464 | 0,001257335 | 0,04166486 | Vibrio | Vibrio metschnikovii |
| 28199 | 28,48006505 | -0,72004793 | 0,198422398 | 12,99024506 | 0,000313118 | 0,024494176 | Arcobacter | Arcobacter nitrofigilis |
| 287 | 749,4267158 | -2,21173488 | 0,373000733 | 32,20684539 | 1,39E-08 | 4,07E-05 | Pseudomonas | Pseudomonas aeruginosa |
| 28885 | 16,19718169 | -0,81108769 | 0,227060435 | 12,21725023 | 0,000473497 | 0,027761739 | Hydrogenovibi | Hydrogenovibrio marinus |
| 29394 | 34,93943003 | -0,40678422 | 0,116742131 | 12,01519342 | 0,000527686 | 0,028681685 | Dolosigranulu | Dolosigranulum pigrum |
| 29433 | 11,87324306 | -1,80115805 | 0,525825147 | 9,933161719 | 0,001623275 | 0,046349306 | Moraxella | Moraxella ovis |
| 296842 | 42,57052307 | 0,368788956 | 0,106995478 | 11,75328953 | 0,000607358 | 0,029963805 | Desulfovibrio | Desulfovibrio carbinolicus |
| 298657 | 39,47459277 | -1,44103937 | 0,278602664 | 26,68661959 | 2,39E-07 | 0,000305216 | Pseudoalteror | Pseudoalteromonas spongiae |
| 300825 | 29,83433421 | -0,65042524 | 0,165442544 | 15,86754954 | 6,79E-05 | 0,012342481 | Alkalihalobaci | Alkalihalobacillus lehensis |
| 308354 | 33,8602557 | -0,66326967 | 0,200786159 | 10,81412428 | 0,001007287 | 0,038375003 | Staphylococcı | Staphylococcus simiae |
| 313588 | 31,43314885 | -0,84372542 | 0,227629429 | 13,91306291 | 0,000191463 | 0,018721908 | Croceibacter | Croceibacter atlanticus |
| 313590 | 13,93132219 | -1,3485571 | 0,323776983 | 17,63547337 | 2,68E-05 | 0,006540507 | Dokdonia | Dokdonia sp. MED134 |
| 313603 | 40,91293764 | -0,78320795 | 0,23593905 | 10,45368395 | 0,001224051 | 0,041629968 | Maribacter | Maribacter sp. HTCC2170 |
| 317658 | 23,18066064 | -0,78245424 | 0,232773221 | 11,20897511 | 0,000814027 | 0,034921856 | Sulfurimonas | Sulfurimonas paralvinellae |
| 326319 | 15,03808259 | -1,05426363 | 0,30215178 | 12,15741343 | 0,000488931 | 0,027761739 | Dokdonia | NA |
| 328515 | 36,29214913 | -0,7602143 | 0,232913329 | 10,41745364 | 0,001248299 | 0,04166486 | Nonlabens | Nonlabens dokdonensis |
| 328813 | 39033,06001 | 1,393906722 | 0,371286677 | 10,88670693 | 0,000968569 | 0,037883954 | Alistipes | Alistipes onderdonkii |
| 33002 | 22,4715117 | -0,79488553 | 0,232191587 | 11,86647485 | 0,000571535 | 0,029674295 | Desulfurella | Desulfurella acetivorans |
| 333965 | 16,54540196 | -0,77617822 | 0,235034428 | 10,98042285 | 0,000920794 | 0,036622114 | Providencia | Providencia vermicola |
| 336810 | 12,53046132 | -0,69638147 | 0,213760023 | 10,19652351 | 0,001407057 | 0,043220949 | Candidatus Su | Candidatus Sulcia muelleri |
| 36818 | 30,99975833 | 0,585079311 | 0,180255697 | 9,760873353 | 0,001782652 | 0,049334055 | Streptomyces | Streptomyces subrutilus |
| 376805 | 8436,30994 | -1,252126 | 0,316737849 | 15,61082855 | 7,78E-05 | 0,012342481 | Phocaeicola | Phocaeicola salanitronis |
| 383750 | 22,0006524 | -0,68058382 | 0,207549275 | 10,71333254 | 0,001063663 | 0,039496894 | Marinomonas | Marinomonas arctica |
| 395922 | 24,92736876 | 0,674071454 | 0,205257028 | 10,30396765 | 0,001327445 | 0,042072712 | Coraliomargar | Coraliomargarita akajimensis |
| 39688 | 27,00780285 | 0,433211401 | 0,140702533 | 9,635582875 | 0,001908436 | 0,049543345 | Mycolicibacte | Mycolicibacterium duvalii |
| 431057 | 7,276379278 | -1,18382415 | 0,369265248 | 10,60714956 | 0,001126512 | 0,040067538 | Methylacidiph | Methylacidiphilum kamchatkense |
| 433296 | 182,3928484 | 0,312478678 | 0,099636608 | 9,882484322 | 0,001668595 | 0,04669 | Aminipila | Aminipila butyrica |
| 43662 | 17,80618588 | -0,68749795 | 0,208118398 | 11,20691813 | 0,000814929 | 0,034921856 | Pseudoalteror | Pseudoalteromonas piscicida |
| 43675 | 138,648101 | 1,12225789 | 0,326669134 | 12,38220975 | 0,000433444 | 0,02764147 | Rothia | Rothia mucilaginosa |
| 44574 | 18,2323443 | -0,52736958 | 0,16609447 | 10,04742182 | 0,001525613 | 0,045205918 | Nitrosomonas | Nitrosomonas communis |

| | | | | | | | | |
|---|---|---|---|---|---|---|---|---|
| 45243 | 66,42456964 | -0,81065574 | 0,245053988 | 10,53144057 | 0,001173607 | 0,040958918 | Capnocytopha | Capnocytophaga haemolytica |
| 47466 | 24,57892189 | -0,80379836 | 0,24891416 | 10,14571713 | 0,001446369 | 0,043968115 | Borrelia | Borrelia miyamotoi |
| 477976 | 43,51200188 | -0,84346982 | 0,222223231 | 13,79733246 | 0,000203625 | 0,01926885 | Calditerrivibri | Calditerrivibrio nitroreducens |
| 47853 | 28,28453287 | 0,450067237 | 0,122726654 | 13,53589149 | 0,000234044 | 0,020494585 | Micromonosp | Micromonospora carbonacea |
| 480520 | 20,18907 | -1,16629497 | 0,256386968 | 20,84491193 | 4,98E-06 | 0,002247573 | Myroides | Myroides profundi |
| 4956 | 31,17231281 | -0,78857451 | 0,204065097 | 14,94029684 | 0,000110967 | 0,014056218 | Zygosaccharo | Zygosaccharomyces rouxii |
| 520734 | 5,976658274 | -1,80092031 | 0,529676239 | 11,41758088 | 0,000727524 | 0,034122543 | Sphingobium | Sphingobium sp. RSMS |
| 52443 | 25,34464995 | -0,91109397 | 0,241989272 | 14,00500639 | 0,000182325 | 0,018443069 | Vibrio | Vibrio tapetis |
| 52773 | 30,17616054 | 0,575944295 | 0,170438158 | 10,67626223 | 0,001085191 | 0,039631706 | Schaalia | Schaalia meyeri |
| 536441 | 77,89503165 | -0,58330126 | 0,18872676 | 9,710630785 | 0,001832051 | 0,049543345 | Planobacteriu | Planobacterium Chryseobacterium taklimakanense |
| 544580 | 5,469151711 | 0,810118548 | 0,257277394 | 9,928489582 | 0,0016274 | 0,046349306 | Actinomyces | Actinomyces oris |
| 545 | 16,46942678 | 1,163823082 | 0,265495748 | 17,84497771 | 2,40E-05 | 0,006535581 | Citrobacter | Citrobacter koseri |
| 561 | 488,5587228 | 1,98373788 | 0,460623426 | 15,33478008 | 9,00E-05 | 0,012884991 | Escherichia | NA |
| 562 | 4903,338991 | 2,103600991 | 0,449144217 | 17,80093525 | 2,45E-05 | 0,006535581 | Escherichia | Escherichia coli |
| 573600 | 9,567738165 | -0,82141114 | 0,229856325 | 12,46751706 | 0,00041409 | 0,027297381 | Candidatus Pl | Candidatus Planktophila limnetica |
| 583 | 31,33004111 | -0,66986693 | 0,204297949 | 11,01737172 | 0,00090262 | 0,036580743 | Proteus | NA |
| 5855 | 55,23893928 | -0,50050392 | 0,161276329 | 9,639786465 | 0,001904074 | 0,049543345 | Plasmodium | Plasmodium vivax |
| 60920 | 26,43022703 | 0,587346976 | 0,153887776 | 14,76874767 | 0,000121533 | 0,014056218 | Sanguibacter | Sanguibacter keddieii |
| 61015 | 25,51839763 | -0,70595147 | 0,2057102 | 12,11314855 | 0,000500675 | 0,027782975 | Staphylococc | Staphylococcus succinus |
| 61435 | 139,0310858 | 0,642710297 | 0,172116675 | 14,24423423 | 0,000160552 | 0,01738733 | Dehalococcoi | Dehalococcoides mccartyi |
| 620 | 13,58911541 | 2,189444541 | 0,563669185 | 14,72202932 | 0,000124582 | 0,014056218 | Shigella | NA |
| 628 | 22,04216551 | -0,79920275 | 0,223371505 | 12,51535087 | 0,000403622 | 0,027297381 | Xenorhabdus | Xenorhabdus nematophila |
| 631220 | 9,243035113 | 0,994417906 | 0,262464495 | 14,73048887 | 0,000124024 | 0,014056218 | Desulfovibrio | Desulfovibrio sp. G11 |
| 645517 | 20,32240347 | 0,465392284 | 0,134425307 | 11,75207635 | 0,000607754 | 0,029963805 | Altererythroba | Altererythrobacter namhicola |
| 668 | 33,77926964 | -0,53968168 | 0,156932025 | 11,62688153 | 0,000650054 | 0,031007071 | Aliivibrio | Aliivibrio fischeri |
| 674 | 28,19856993 | -0,65677091 | 0,192194596 | 11,78046132 | 0,000598556 | 0,029963805 | Vibrio | Vibrio mimicus |
| 68223 | 27,64789616 | 0,53058609 | 0,149242713 | 12,48761449 | 0,000409659 | 0,027297381 | Streptomyces | Streptomyces katrae |
| 68288 | 9,037544856 | -1,70175812 | 0,479875944 | 10,62180357 | 0,00111762 | 0,040067538 | Cyclobacteriu | NA |
| 683124 | 32,28158119 | -0,57736662 | 0,187420273 | 9,675419356 | 0,001867496 | 0,049543345 | Flavobacteriu | Flavobacterium haoranii |
| 698776 | 193,2343054 | -0,3490778 | 0,107679494 | 10,5216686 | 0,001179829 | 0,040958918 | Cellulosilyticu | NA |
| 69966 | 31,99987624 | -0,7294521 | 0,183802368 | 15,72216864 | 7,34E-05 | 0,012342481 | Macrococcus | Macrococcus caseolyticus |
| 712116 | 16,89683499 | 0,730469422 | 0,223420631 | 9,696227736 | 0,001846467 | 0,049543345 | Actinomyces | Actinomyces sp. oral taxon 169 |
| 712122 | 35,39261981 | 0,411130268 | 0,12914606 | 10,04928044 | 0,001524075 | 0,045205918 | Actinomyces | Actinomyces sp. oral taxon 414 |
| 712623 | 63,75362387 | 1,028233093 | 0,304367055 | 11,39513549 | 0,000736367 | 0,034122543 | Streptococcus | Streptococcus sp. oral taxon 061 |
| 712633 | 44,278314 | 1,108496445 | 0,332717452 | 11,37485701 | 0,00074445 | 0,034122543 | Streptococcus | Streptococcus sp. oral taxon 431 |
| 713887 | 22,96612056 | -0,90531447 | 0,247421364 | 13,75891637 | 0,000207832 | 0,019354764 | Candidatus At | Candidatus Atelocyanobacterium thalassa |
| 758 | 18,57422338 | -0,75879845 | 0,223584236 | 11,38160575 | 0,00074175 | 0,034122543 | Rodentibacter | Rodentibacter pneumotropicus |
| 758802 | 36,11050824 | 0,432066514 | 0,121350803 | 12,75265573 | 0,000355504 | 0,026071777 | Mycolicibacte | Mycolicibacterium litorale |
| 762954 | 26,10047026 | -0,69279649 | 0,209984297 | 10,85765637 | 0,000983881 | 0,038047249 | Siansivirga | Siansivirga zeaxanthinifaciens |
| 76594 | 46,38006811 | -0,83090939 | 0,222416498 | 13,79928878 | 0,000203413 | 0,01926885 | Cellulophaga | Cellulophaga baltica |
| 782 | 4,165730335 | 1,109059454 | 0,329400922 | 11,78882684 | 0,000595873 | 0,029963805 | Rickettsia | Rickettsia prowazekii |
| 78543 | 7,998113896 | 0,750104817 | 0,217385508 | 11,77936039 | 0,000598911 | 0,029963805 | Pseudomonas | Pseudomonas migulae |
| 81462 | 25,50056198 | 0,51544897 | 0,148245995 | 12,21553501 | 0,000473932 | 0,027761739 | Thermanaerov | Thermanaerovibrio acidaminovorans |
| 824 | 20,75024808 | 0,980624513 | 0,254583837 | 14,94516624 | 0,000110681 | 0,014056218 | Campylobacte | Campylobacter gracilis |
| 830 | 218,1587517 | -0,5195391 | 0,15815506 | 10,59544714 | 0,001133665 | 0,040067538 | Butyrivibrio | NA |
| 84112 | 512,6282863 | 1,263612896 | 0,291597193 | 15,78236223 | 7,11E-05 | 0,012342481 | Eggerthella | Eggerthella lenta |
| 85471 | 100,0025282 | -0,42616666 | 0,131726173 | 10,46975724 | 0,001213447 | 0,041629968 | Plasmodium | Plasmodium relictum |
| 912630 | 22,22041755 | -0,55785893 | 0,182974585 | 9,685099591 | 0,001857683 | 0,049543345 | Microbacteriu | Microbacterium sp. LKL04 |
| 935222 | 35,97936606 | -0,73701405 | 0,19460212 | 14,76773415 | 0,000121598 | 0,014056218 | Flavobacteriu | Flavobacterium sp. Sr18 |
| 97084 | 58,24785602 | -1,02295044 | 0,296557354 | 11,70896421 | 0,000621997 | 0,030198474 | Halobacteriov | Halobacteriovorax marinus |
| 97393 | 4,700875136 | -1,63426761 | 0,421185792 | 15,45330697 | 8,46E-05 | 0,012722189 | Ferroplasma | Ferroplasma acidarmanus |
| 985 | 52,45385908 | -0,51044385 | 0,161444552 | 10,06509686 | 0,001511046 | 0,045205918 | Cytophaga | Cytophaga hutchinsonii |
| 985762 | 22,43792033 | -0,68833856 | 0,193413552 | 12,48618014 | 0,000409974 | 0,027297381 | Staphylococc | Staphylococcus agnetis |
| 996 | 97,92724171 | -0,72932116 | 0,206005217 | 12,6059984 | 0,000384511 | 0,027297381 | Flavobacteriu | Flavobacterium columnare |
