## Supplementary material for "Gut microbiota dysbiosis in adrenoleukodystrophy: a new piece in the puzzle?": ES2_functions_general_sequential_model

| baseMean | log2FoldChan | lfcSE | stat | pvalue | padj | Subsystem3 |
|---|---|---|---|---|---|---|
| 22,6832092 | 2,22106889 | 0,39381266 | 32,7278041 | 1,06E-08 | 9,66E-06 | Mediator of hyperadherence YidE in Enterobacteria and its conserved region |
| 16,6152268 | 3,85963102 | 0,66288464 | 30,7089102 | 3,00E-08 | 1,37E-05 | Colonization factor antigen I fimbriae |
| 73,3482065 | 1,13616054 | 0,20686705 | 29,4373505 | 5,78E-08 | 1,75E-05 | D-Sorbitol(D-Glucitol) and L-Sorbose Utilization |
| 7,31639572 | 2,30051888 | 0,43742648 | 28,1872632 | 1,10E-07 | 2,51E-05 | Commensurate regulon activation |
| 752,46849 | -0,24988766 | 0,04779768 | 27,4153786 | 1,64E-07 | 2,99E-05 | At3g21300 |
| 77,4832945 | 1,72674433 | 0,36211113 | 23,57256 | 1,20E-06 | 0,00013698 | Type 4 secretion and conjugative transfer |
| 61,132087 | 1,14980695 | 0,23656948 | 24,0624359 | 9,33E-07 | 0,00013698 | Unknown carbohydrate utilization ( cluster Yeg ) |
| 14,7294463 | 1,54178106 | 0,32527505 | 23,7113316 | 1,12E-06 | 0,00013698 | Glyoxylate bypass |
| 6,36473663 | 2,02077121 | 0,4284782 | 22,8204309 | 1,78E-06 | 0,00018004 | Biogenesis of cytochrome c oxidases |
| 17,3004509 | 1,50763941 | 0,31117627 | 22,4911512 | 2,11E-06 | 0,00019232 | Siderophore assembly kit |
| 158,486499 | 0,97911881 | 0,20876874 | 22,0485625 | 2,66E-06 | 0,00022016 | Orphan regulatory proteins |
| 13,2496758 | 1,57384838 | 0,34008934 | 21,3822972 | 3,76E-06 | 0,00028562 | Ferrous iron transporter EfeUOB, low-pH-induced |
| 4,59134719 | 1,49756526 | 0,32825166 | 21,1387758 | 4,27E-06 | 0,00029937 | Small multidrug resistance strays |
| 138,708162 | 0,95936378 | 0,20658206 | 20,7706074 | 5,18E-06 | 0,00033688 | Unknown carbohydrate utilization ( cluster Ydj ) |
| 29,7586977 | 1,04874641 | 0,2336314 | 20,6234532 | 5,59E-06 | 0,00033954 | Regulation of Oxidative Stress Response |
| 14,3219142 | 1,87674265 | 0,41502072 | 20,4762887 | 6,04E-06 | 0,00034376 | D-galactonate catabolism |
| 22,753889 | 1,81614149 | 0,40223511 | 20,0695364 | 7,47E-06 | 0,00037795 | Two partner secretion pathway (TPS) |
| 28,5264013 | 1,342867 | 0,30865356 | 20,1139227 | 7,30E-06 | 0,00037795 | Putrescine utilization pathways |
| 17,7723876 | 1,29790086 | 0,30241916 | 19,5303062 | 9,90E-06 | 0,00047476 | Universal stress protein family |
| 24,3875352 | 2,04059173 | 0,43596061 | 19,2985225 | 1,12E-05 | 0,00050922 | Carnitine Metabolism in Microorganisms |
| 183,174913 | 0,47254288 | 0,10640558 | 19,0670077 | 1,26E-05 | 0,0005475 | Mannitol Utilization |
| 13,7837725 | 1,24587578 | 0,29268641 | 18,9188795 | 1,36E-05 | 0,0005648 | A Gammaproteobacteria Cluster Relating to Translation |
| 49,6078402 | 1,27119393 | 0,29907988 | 18,1559714 | 2,04E-05 | 0,00079275 | Lipid A modifications |
| 18,4127576 | 2,19510369 | 0,46388629 | 18,1068567 | 2,09E-05 | 0,00079275 | Siderophore Yersiniabactin Biosynthesis |
| 14,8921957 | 1,25990189 | 0,30140554 | 17,7532634 | 2,51E-05 | 0,00091641 | Phosphoglycerate transport system |
| 50,49115 | 0,85353361 | 0,20052163 | 17,6457198 | 2,66E-05 | 0,00093242 | Tolerance to colicin E2 |
| 15,2017376 | 1,24288445 | 0,3043473 | 17,4042907 | 3,02E-05 | 0,00101945 | Late competence |
| 2882,10301 | -0,13821991 | 0,03308786 | 17,3302544 | 3,14E-05 | 0,00102209 | Translation termination factors bacterial |
| 677,63795 | -0,18842009 | 0,04578168 | 16,9952114 | 3,75E-05 | 0,00114579 | Glycine cleavage system |
| 3,7881777 | 1,63559627 | 0,40490051 | 16,9821996 | 3,77E-05 | 0,00114579 | Single-Rhodanese-domain proteins |
| 8,7957631 | 1,9192875 | 0,4690263 | 16,5907389 | 4,64E-05 | 0,00136288 | Tellurite resistance: Chromosomal determinants |
| 31,774243 | 2,45434284 | 0,61568429 | 16,2613077 | 5,52E-05 | 0,00156871 | Type 1 pili (mannose-sensitive fimbriae) |
| 5,33240617 | 1,61562997 | 0,40821173 | 16,2056387 | 5,68E-05 | 0,00156871 | CBSS-211586.1.peg.3133 |
| 4,07188593 | 1,95430935 | 0,49815187 | 15,9299135 | 6,57E-05 | 0,00176122 | CytR regulation |
| 6,72084981 | 2,51432219 | 0,59681724 | 15,8409404 | 6,89E-05 | 0,00179325 | CBSS-194948.1.peg.143 |
| 802,501134 | -0,14542507 | 0,03682463 | 15,5432922 | 8,06E-05 | 0,00204057 | Ribosomal protein S12p Asp methylthiotransferase |
| 105,668663 | 0,66866829 | 0,1700017 | 15,2291832 | 9,52E-05 | 0,00234448 | Putative sugar ABC transporter (ytf cluster) |
| 4,27896041 | 1,96002011 | 0,51909427 | 15,032389 | 0,00010568 | 0,00252037 | Intracellular septation in Enterobacteria |
| 12,910037 | 0,92850287 | 0,23836427 | 14,9932349 | 0,0001079 | 0,00252037 | Molybdopterin cytosine dinucleotide |
| 11,4799348 | 1,11201378 | 0,29445019 | 14,6777137 | 0,00012755 | 0,00290485 | Streptococcus pyogenes Virulome |
| 15,0286711 | 2,14361417 | 0,54677437 | 14,6054441 | 0,00013253 | 0,00294478 | Biofilm Adhesin Biosynthesis |
| 1,17331475 | 1,80488925 | 0,48205344 | 14,478758 | 0,00014175 | 0,00305951 | CBSS-196620.1.peg.2477 |
| 37,2768914 | 0,67639596 | 0,17335453 | 14,4403114 | 0,00014467 | 0,00305951 | RNA 3'-terminal phosphate cyclase |
| 1,4637512 | 2,02477702 | 0,54581065 | 14,4004098 | 0,00014777 | 0,00305951 | RNA processing orphans |
| 25,5921084 | 0,70425321 | 0,18892834 | 14,3009215 | 0,00015579 | 0,00315385 | Putative oxidase COG2907 |
| 2,80075583 | 1,83857573 | 0,50152848 | 13,9408546 | 0,00018865 | 0,00369251 | CBSS-269801.1.peg.2186 |
| 29,7843834 | 1,36386235 | 0,37642254 | 13,9225136 | 0,0001905 | 0,00369251 | Formate dehydrogenase |
| 1241,14805 | -0,21689528 | 0,05850568 | 13,8057346 | 0,00020272 | 0,00384739 | Cysteine Biosynthesis, MCB 432 |
| 10,1508717 | 1,48917244 | 0,38335967 | 13,6393539 | 0,00022149 | 0,00411797 | carbazol degradation cluster |
| 927,302224 | -0,17387639 | 0,04697053 | 13,5702919 | 0,00022979 | 0,00418684 | TenI-like tautomerase |
| 4779,59536 | -0,09569389 | 0,02596496 | 13,5158681 | 0,00023655 | 0,00422552 | Heat shock dnaK gene cluster extended |
| 2119,44022 | -0,12205966 | 0,03311732 | 13,446736 | 0,00024543 | 0,00429979 | tRNA aminoacylation, Phe |
| 3779,07005 | -0,13666459 | 0,03743658 | 13,2433374 | 0,00027355 | 0,00470197 | Bacterial Cell Division |
| 3,88885103 | 2,06617654 | 0,58990557 | 13,0933937 | 0,00029634 | 0,0049744 | Unknown carbohydrate utilization containing Fructose-bisphosphate aldolase |
| 18,4593585 | 0,81017505 | 0,22773271 | 13,0683938 | 0,00030032 | 0,0049744 | CBSS-211586.1.peg.2357 |
| 3826,07355 | -0,12560687 | 0,03477839 | 12,9658665 | 0,00031722 | 0,00516052 | RP Bacterial Cell Division |
| 21,9357747 | 1,085971 | 0,31043578 | 12,824109 | 0,00034218 | 0,0054689 | Beta-lactamase cluster in Streptococcus |
| 2406,94535 | -0,130363 | 0,03640529 | 12,7214674 | 0,00036148 | 0,00558153 | tRNA aminoacylation, Glu and Gln |
| 496,091652 | -0,21885907 | 0,06114156 | 12,7360638 | 0,00035867 | 0,00558153 | Mycobacterium virulence operon possibly involved in quinolinate biosynthesis |
| 3,48777409 | 2,01830242 | 0,52922476 | 12,6358078 | 0,00037843 | 0,00571769 | Capsular Polysaccharide (CPS) of Campylobacter |
| 21,5059556 | 0,55912186 | 0,15967714 | 12,6140756 | 0,00038285 | 0,00571769 | Adaptation to d-cysteine |
| 268,419782 | 0,36971251 | 0,10523995 | 12,4852988 | 0,00041017 | 0,00598215 | Type VI secretion systems |
| 742,324125 | -0,47412833 | 0,13494918 | 12,4693037 | 0,00041369 | 0,00598215 | Flagellar motility |
| 599,386757 | -0,15783901 | 0,04474211 | 12,3949394 | 0,0004305 | 0,00612788 | Serine Biosynthesis |
| 3055,75324 | -0,16928254 | 0,04808956 | 12,2408562 | 0,00046754 | 0,0065528 | F0F1-type ATP synthase |
| 136,972178 | 0,60947018 | 0,17700038 | 11,9859457 | 0,00053603 | 0,00739888 | Ubiquinone Biosynthesis |
| 13,449929 | 1,19237713 | 0,35382639 | 11,7589661 | 0,00060551 | 0,00823311 | SeqA and Co-occurring Genes |
| 4,9448382 | 1,03719032 | 0,30652097 | 11,699928 | 0,00062503 | 0,0083735 | Coenzyme PQQ synthesis |
| 12,0823748 | 2,18584281 | 0,61525442 | 11,6646128 | 0,000637 | 0,00841027 | The fimbrial Stf cluster |
| 18,0746722 | 1,18286665 | 0,35824434 | 11,394602 | 0,00073658 | 0,00958604 | Competence in Streptococci |
| 128,243559 | 0,62958626 | 0,19098495 | 11,307158 | 0,00077209 | 0,00990666 | Lipopolysaccharide assembly |
| 31,4266366 | 1,09626959 | 0,33744839 | 11,2543909 | 0,00079435 | 0,01005072 | Phospholipid and Fatty acid biosynthesis related cluster |
| 4673,08731 | -0,12145742 | 0,03611639 | 11,21533 | 0,00081124 | 0,01012388 | Single-copy ribosomal proteins |
| 2380,59097 | -0,08803267 | 0,02630656 | 11,1607863 | 0,00083545 | 0,01028501 | Competence or DNA damage-inducible protein CinA and related protein families |
| 2,35046432 | 3,57180185 | 1,06729325 | 10,959209 | 0,0009314 | 0,01116449 | β-Fimbriae |
| 9,24375359 | 2,37537564 | 0,67044624 | 10,9814393 | 0,00092029 | 0,01116449 | Lysozyme inhibitors |
| 284,013981 | 0,86298211 | 0,24729562 | 10,7527149 | 0,00104126 | 0,01231938 | Phd-Doc, YdcE-YdcD toxin-antitoxin (programmed cell death) systems |
| 2124,27599 | -0,11333873 | 0,03459408 | 10,7180089 | 0,00106098 | 0,01239168 | UDP-N-acetylmuramate from Fructose-6-phosphate Biosynthesis |
| 14,0241213 | 0,83181732 | 0,26397071 | 10,5185763 | 0,0011818 | 0,01362816 | Benzoate degradation |
| 5840,40309 | -0,10593221 | 0,03263765 | 10,4589063 | 0,0012206 | 0,01389953 | DNA repair, UvrABC system |
| 603,569914 | -0,1657556 | 0,05135256 | 10,3285419 | 0,00130989 | 0,01473218 | Methylthiotransferases |

| | | | | | | |
|---|---|---|---|---|---|---|
| 5821,36697 | -0,10386684 | 0,03230224 | 10,2871816 | 0,00133958 | 0,01488236 | Glutamine, Glutamate, Aspartate and Asparagine Biosynthesis |
| 2,03295078 | 1,67049649 | 0,54398313 | 10,09322 | 0,00148816 | 0,01613943 | Iron acquisition in Vibrio |
| 15,6651681 | 0,72705497 | 0,22172913 | 10,1096733 | 0,00147493 | 0,01613943 | Tagatose utilization |
| 9,92803298 | 1,58245969 | 0,51083795 | 10,0261493 | 0,00154333 | 0,0165409 | Curli production |
| 1669,31448 | -0,1281883 | 0,04051282 | 9,99504583 | 0,00156962 | 0,01662701 | tRNA aminoacylation, Ile |
| 6203,55665 | -0,17600635 | 0,05559422 | 9,9116674 | 0,00164234 | 0,01707944 | Glycogen metabolism |
| 4367,24029 | -0,11794848 | 0,03730732 | 9,90330117 | 0,00164983 | 0,01707944 | Methionine Biosynthesis |
| 63,5727684 | 0,4894595 | 0,15935312 | 9,87339871 | 0,00167685 | 0,01716421 | Autoinducer 2 (AI-2) transport and processing (lsrACDBFGE operon) |
| 288,056534 | -0,18859898 | 0,0605473 | 9,69405706 | 0,00184865 | 0,01850681 | Adenosyl nucleosidases |
| 119,644022 | 0,46997891 | 0,15191418 | 9,70494332 | 0,00183773 | 0,01850681 | Lysine degradation |
| 2,60680475 | 2,46296283 | 0,79923975 | 9,58531461 | 0,0019614 | 0,01900887 | P uptake (cyanobacteria) |
| 1299,23417 | -0,06789874 | 0,02186742 | 9,60900893 | 0,00193625 | 0,01900887 | Pyridoxin (Vitamin B6) Biosynthesis |
| 4,69379573 | 1,25895112 | 0,42141899 | 9,59719059 | 0,00194875 | 0,01900887 | Quorum sensing in Yersinia |
| 1900,95881 | -0,16556551 | 0,0540076 | 9,35984772 | 0,00221791 | 0,02126855 | CarD |
| 17,9319541 | 0,53662611 | 0,17645497 | 9,33699201 | 0,00224574 | 0,02131112 | Khodge314 Isoleucine Biosynthesis |
| 3985,67364 | -0,0885649 | 0,0292054 | 9,15553338 | 0,00247967 | 0,02305082 | Transcription factors bacterial |
| 131,671991 | 0,77316764 | 0,24497771 | 9,16834957 | 0,00246237 | 0,02305082 | Integrons |
| 1168,98073 | -0,14140833 | 0,0467482 | 9,11863674 | 0,00253018 | 0,02328281 | coA-FolK |
| 6170,65014 | -0,11870516 | 0,03921735 | 9,09406431 | 0,0025644 | 0,02336172 | Universal GTPases |
| 32,735259 | 0,86333401 | 0,28598697 | 8,86941347 | 0,00289988 | 0,02615637 | trimethylamine N-oxide (TMAO) reductase |
| 1119,66445 | -0,14123096 | 0,04766523 | 8,66291944 | 0,00324751 | 0,02892173 | Leucine Biosynthesis |
| 3,28718879 | 0,83187867 | 0,28707292 | 8,65036144 | 0,00326996 | 0,02892173 | Nitric oxide synthase |
| 40,4742968 | 0,52930838 | 0,18046603 | 8,61623801 | 0,00333179 | 0,02918522 | Fructose and Mannose Inducible PTS |
| 2,12800297 | 0,88515062 | 0,30226555 | 8,55266761 | 0,00345016 | 0,02993427 | Sulfur oxidation |
| 3044,42613 | -0,07628451 | 0,02611494 | 8,46105438 | 0,00362831 | 0,03118297 | Protein chaperones |
| 976,70991 | -0,14512704 | 0,04987844 | 8,4427695 | 0,00366498 | 0,03120368 | tRNA aminoacylation, Arg |
| 66,4598228 | 0,32128088 | 0,11221325 | 8,34366296 | 0,00387035 | 0,03234759 | EC 3.4.19.- Omega peptidases |
| 4781,45175 | -0,14393503 | 0,04956664 | 8,34543334 | 0,00386658 | 0,03234759 | Glycolysis and Gluconeogenesis |
| 489,45525 | -0,08814429 | 0,0306178 | 8,24783967 | 0,00408005 | 0,03379026 | Glutamine synthetases |
| 3329,30535 | -0,13881421 | 0,04838985 | 8,11456747 | 0,00439109 | 0,03450977 | Branched-Chain Amino Acid Biosynthesis |
| 19,3958216 | 0,61773484 | 0,21541626 | 8,10793715 | 0,00440719 | 0,03450977 | Citrate Metabolism KE2 |
| 5751,39148 | -0,11213774 | 0,0392594 | 8,10422983 | 0,00441621 | 0,03450977 | De Novo Purine Biosynthesis |
| 42,5912204 | 0,46739367 | 0,16509416 | 8,16164921 | 0,00427854 | 0,03450977 | Homogentisate pathway of aromatic compound degradation |
| 79,4710545 | 0,35376052 | 0,12460544 | 8,1499325 | 0,00430628 | 0,03450977 | L-Cystine Uptake and Metabolism |
| 555,639783 | -0,11135864 | 0,03907759 | 8,09771898 | 0,0044321 | 0,03450977 | Ribonuclease H |
| 2,32327121 | 1,45558478 | 0,50527057 | 8,18409789 | 0,00422592 | 0,03450977 | Streptococcal Mga Regulon |
| 3490,62586 | -0,11477696 | 0,04033198 | 8,05028667 | 0,00454965 | 0,03482964 | Ribosome biogenesis bacterial |
| 376,439246 | -0,25624898 | 0,09014407 | 8,0620528 | 0,00452019 | 0,03482964 | Sporulation-associated proteins with broader functions |
| 132,989193 | -0,14476627 | 0,0511431 | 8,01665783 | 0,0046349 | 0,03518664 | t(6)A synthesis in bacteria |
| 1,33113995 | 1,11806095 | 0,40942702 | 7,84146887 | 0,00510611 | 0,03844352 | Alpha-acetolactate operon |
| 872,839861 | -0,18420901 | 0,06710926 | 7,48142601 | 0,00623387 | 0,04423664 | dcernst CoA Salvage |
| 2024,84946 | -0,12534368 | 0,04546463 | 7,55997527 | 0,00596788 | 0,04423664 | tRNA aminoacylation, Asp and Asn |
| 8,06454822 | 0,52219767 | 0,18829146 | 7,5009472 | 0,00616666 | 0,04423664 | Carotenoids |
| 4151,20469 | -0,09408111 | 0,03428913 | 7,50784818 | 0,00614307 | 0,04423664 | Folate Biosynthesis |
| 7,51472424 | 0,5350845 | 0,19561702 | 7,54348384 | 0,00602275 | 0,04423664 | Osmotic stress cluster |
| 1159,1184 | -0,09361276 | 0,03400214 | 7,54950953 | 0,00600264 | 0,04423664 | Polyadenylation bacterial |
| 657,440034 | -0,14691795 | 0,05341053 | 7,47273699 | 0,00626403 | 0,04423664 | Proline Synthesis |
| 4763,7645 | -0,09447731 | 0,03440796 | 7,4827926 | 0,00622914 | 0,04423664 | Proteolysis in bacteria, ATP-dependent |
| 745,538566 | -0,11467747 | 0,04182348 | 7,4176442 | 0,00645873 | 0,04440552 | tRNA aminoacylation, Trp |
| 0,97429898 | 1,30189709 | 0,50414203 | 7,3974453 | 0,00653166 | 0,04440552 | Chlorobenzoate degradation |
| 30,4401879 | 0,33129658 | 0,12321332 | 7,40842292 | 0,00649192 | 0,04440552 | Dissimilatory nitrite reductase |
| 2,47627416 | 1,44834961 | 0,55159977 | 7,40927804 | 0,00648884 | 0,04440552 | Selenoprotein O |
| 2774,53836 | -0,12813771 | 0,04697243 | 7,42542109 | 0,00643088 | 0,04440552 | At5g63290 |
| 255,44726 | 0,3885961 | 0,14395669 | 7,31897281 | 0,00682304 | 0,04537072 | Transport of Nickel and Cobalt |
| 353,847544 | 0,38367684 | 0,14162582 | 7,33301871 | 0,00676993 | 0,04537072 | Nitrate and nitrite ammonification |
| 949,021482 | -0,12814325 | 0,04717315 | 7,31953156 | 0,00682092 | 0,04537072 | NusA-TFII Cluster |
| 119,108195 | 0,39953214 | 0,14867636 | 7,30145004 | 0,0068899 | 0,04548332 | Bacterial hemoglobins |
| 1930,61948 | -0,12246691 | 0,04546426 | 7,26477627 | 0,007032 | 0,04608744 | tRNA processing |
| 648,730074 | -0,10440731 | 0,038841 | 7,20181449 | 0,00728299 | 0,04739146 | Alanine biosynthesis |
| 591,606325 | -0,14709649 | 0,05496517 | 7,16355624 | 0,00743995 | 0,0480695 | tRNA nucleotidyltransferase |
| 56,0638427 | 0,48214664 | 0,17744871 | 7,15078882 | 0,0074931 | 0,04807195 | Glycerol fermentation to 1,3-propanediol |
| 1229,25726 | -0,08726352 | 0,03262723 | 7,13075731 | 0,00757727 | 0,04827199 | Translation initiation factors bacterial |
| 2863,69882 | -0,10031256 | 0,03762403 | 7,08272139 | 0,00778307 | 0,04923874 | Threonine and Homoserine Biosynthesis |
